# Tracking community infection dynamics of COVID-19 by monitoring SARS-CoV-2 RNA in wastewater, counting positive reactions by qPCR

**DOI:** 10.1101/2021.12.23.21268343

**Authors:** Bo Zhao, Zaizhi Yu, Tomonori Fujita, Yoshiaki Nihei, Hiroaki Tanaka, Masaru Ihara

**Affiliations:** Research Center for Environmental Quality Management, Graduate School of Engineering, Kyoto University, 1-2 Yumihama, Otsu, Shiga 520-0811, Japan; Water Agency Inc. 3-25 Higashi-Goken-cho, Shinjuku-ku, Tokyo 162-0813, Japan; Faculty of Agriculture and Marine Science, Graduate School of Integrated Arts and Sciences, Kochi University, 200 Otsu, Monobe, Nankoku city, Kochi 783-8502, Japan

**Keywords:** Wastewater-based epidemiology, SARS-CoV-2, Poisson distribution, Positive ratio, qPCR, Low prevalence

## Abstract

Wastewater-based epidemiology has proved useful for monitoring the COVID-19 infection dynamics in communities. However, in some countries, low concentrations of SARS-CoV-2 RNA in wastewater make this difficult. Getting meaningful information from wastewater-based epidemiology in regions of low prevalence remains a key challenge. Here we used real-time reverse-transcription PCR (RT-qPCR) to monitor SARS-CoV-2 RNA in wastewater from October 2020 to February 2021 during the third wave of the COVID-19 outbreak in Japan. Viral RNA was below the limit of quantification in all samples. However, by counting the positive reactions in repeated qPCR of each sample, we found that the ratio of positive reactions to all tests in wastewater was significantly correlated with the number of clinically confirmed cases by the date of symptom onset during periods of both increasing and decreasing infection. Time-step analysis indicated that COVID-19 patients excreted large amounts of virus in their feces 2 days either side of symptom onset, which wastewater surveillance could detect. The positive count method is thus useful for tracing COVID-19 dynamics in regions of low prevalence.

**Highlights:**

- Positive ratio by repeated qPCR of low target-molecule numbers correlated with number expected from Poisson distribution.
- Positive ratio by repeated RT-qPCR of SARS-CoV-2 RNA in wastewater tracked the infection dynamics of COVID-19 in a region of low prevalence.
- Positive ratios correlated with number of new cases by date of symptom onset.
- COVID-19 patients might excrete more virus in their feces in the period from 2 days before to 2 days after symptom onset.

## Introduction

A novel coronavirus—severe acute respiratory syndrome coronavirus 2 (SARS-CoV-2)—was identified in Wuhan in late December 2019,^1^ and has since caused a global pandemic of coronavirus disease 2019 (COVID-19) owing to its fast spread. The progression of the COVID-19 pandemic has been monitored primarily by clinical testing of symptomatic individuals for the presence of the SARS-CoV-2 RNA by polymerase chain reaction (PCR) analysis. However, in many countries, the spread of the virus has exceeded testing capacity, precluding real-time monitoring of the pandemic. Instead, wastewater-based epidemiology (WBE) has been considered as an effective approach for monitoring the presence of SARS-CoV-2 in the community.^2^

Increases and decreases of viral RNA concentrations measured by real-time reverse-transcription PCR (RT-qPCR) in raw influent or its solid fraction have been associated with those of COVID-19 cases in Australia^3^, the Netherlands^4^, the USA^5-8^, and Canada^9,10^. In Japan, some studies have detected SARS-CoV-2 RNA in wastewater when cases were diagnosed by clinical testing in the community.^11-14^ Concentrations in the solid fraction of raw influent were quantifiable,^11^ but the supernatant fraction had high PCR threshold cycle (Ct) values, and most data were unquantifiable owing to low concentrations of SARS-CoV-2 RNA in the wastewater.^12-14^ Getting meaningful information from WBE in regions of low prevalence remains a key challenge.

In general, the limit of quantification (LOQ) of qPCR for the virus in wastewater is based on the fewest copies of the control molecule in a reaction volume that can be quantitatively determined with a stated probability; e.g., 5 or 10 copies per reaction. For wastewater with Ct smaller than those of 5 or 10 copies, the virus RNA copy number is titrated. These wastewater is rated either positive or negative for the virus RNA relative to the LOQ. In qPCR, when the number of initial target molecules is <10, the probability that an aliquot contains a given number of target molecules is given by a Poisson distribution, with which the pattern of positive and negative results is consistent.^15-17^ In this range, the chance of a positive increases with the number of initial target molecules in the solution. Thus, we hypothesized that by calculating the positive ratio (the ratio of positive reactions to all tests) in repeated qPCR analysis of wastewater, we might track the infection dynamics even in regions of low prevalence.

The purpose of this study was to verify the effectiveness of calculating the positive ratio for SARS-CoV-2 RNA in wastewater to track the infection dynamics of COVID-19 in the community. First, we tested the pattern of positive and negative results in qPCR in the range of 0.1 to 20 initial copies of oligo DNA including the CDC-N1 target per reaction, and confirmed that empirical data obtained by qPCR of these samples followed a Poisson distribution. Then we took raw or primary effluent from wastewater treatment plant (WWTP) A in city B in the Kansai region of Japan almost every day from October 2020 to February 2021, during the third wave of COVID-19 in Japan. After polyethylene glycol (PEG) precipitation to concentrate the virus, we analyzed SARS-CoV-2 RNA in the wastewater by RT-qPCR. The positive ratio for SARS-CoV-2 RNA in technically repeated qPCR was compared with epidemiological data in the community reported by the public health system. In most samples, SARS-CoV-2 RNA was below the LOQ, but the positive ratio in repeated qPCR for N1 and N2 assays in wastewater was significantly correlated with the number of daily confirmed cases by the date of symptom onset. Time-step analysis indicates that COVID-19 patients excreted a large amount of virus in their feces during the period from 2 days before to 2 days after symptom onset, which wastewater surveillance could detect.

## Materials and Methods

### Verification of concordance between positive counts in qPCR of low-target-copy-number samples and Poisson distribution

The Poisson distribution for PCR is defined as:^15^

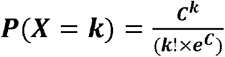

where e is the base of the natural logarithm, *C* is the average initial target molecule number (ITMN) per qPCR sample (expected ITMN), and *k* is the actual number (actual ITMN) in a sample. *P* is the probability that a volume (sample) contains *k* copies of ITMN. SI Figure S1 shows the probabilities of obtaining a certain number of target molecules in a given volume, where the sample average concentration ranges from 10 to 0.01 copies per volume.

qPCR assays for low target copy number were performed. Oligo DNA including the SARS-CoV-2 sequence (Thermo Scientific) was diluted to prepare different ITMN samples (*C* = 0.01, 0.1, 0.2, 0.4, 0.6, 0.8, 1, 1.2, 1.4, 1.6, 1.8, 2, 4, 6, 8, or 10 copies per reaction), and the CDC 2019-nCoV_N1 assay^18^ was performed 12 times for each ITMN sample. Details of the qPCR for SARS-CoV-2 are described below under “qPCR for SARS-CoV-2”. We counted the positives in the 12 qPCR reactions for each ITMN sample, and then compared the resulting “positive ratios” with those expected from the Poison distribution. These experiments were performed three times (three batches).

### Wastewater sample collection

Influent or primary effluent (PE) was collected at WWTP A in city B. PE was collected two times (Tuesday and Friday) a week from 20 October to 14 December 2020, and five times (Monday to Friday) a week from 15 December 2020 to 14 January 2021. Influent was collected five times (Monday to Friday) a week from 15 January to 15 February. In total, 60 samples (39 PE and 21 influent) were collected in the morning (09:00–10:00).

In October 2020, clinical testing in city B confirmed few to no daily COVID-19 cases. From November 2020 to February 2021, city B experienced an increase followed by a decrease in the number of cases, during the third wave of infection in Japan.

Wastewater was collected in a sterilized 250-mL plastic bottle, immediately transported to the laboratory, and then heat-treated in water bath (60 °C, 90 min) to inactivate the coronavirus.^19,20^ The heat-treated samples were stored at −30 °C. Within a week after collection, they were analyzed for SARS-CoV-2 RNA by RT-qPCR.

### Virus concentration in wastewater, RNA extraction, and reverse transcription

Virus in each sample was concentrated by PEG precipitation^13^ with a slight modification. First, 120 mL of PE was centrifuged at 4500× *g* for 10 min, and the supernatant was transferred to a fresh centrifuge tube. Then, PEG 8000 (Molecular Biology Grade, average mol wt 8000; Sigma-Aldrich) and NaCl were added to final concentrations of 10% and 1 M, respectively. The samples were incubated at 4 °C overnight with gentle agitation. After centrifugation at 10□000× *g* for 30 min, the PEG precipitate, containing the virus, was dissolved in 500 μL of phosphate buffer solution (for biochemistry, 0.1 M, pH 8.0, Wako) to give a total volume of ∼700 μL. From 140 μL of the virus concentrate, RNA was extracted with a QIAamp Viral RNA Mini kit (Qiagen, Hilden, Germany) as per the manufacturer’s instructions to obtain an 80-μL RNA extract. For the RNA extraction step, PCR-grade water (Roche Molecular Systems, Inc., Switzerland) was included as a negative control every time.

For reverse transcription (RT), a High-Capacity cDNA Reverse Transcription Kit (Applied Biosystems, Foster City, CA, USA) was used to obtain 70 μL of cDNA from 35 μL of viral RNA as per the manufacturer’s protocol. cDNA was amplified by qPCR to detect SARS-CoV-2 and Pepper Mild Mottle Virus (PMMoV). For the RT step, PCR-grade water was included as a negative control every time.

### qPCR for SARS-CoV-2

The SARS-CoV-2 RNA was assayed by the CDC 2019-nCoV_N1 and CDC 2019-nCoV_N2 qPCR assays^18^ in all samples with the primers and probes listed in Table S1. qPCR assays in 96-well plates were conducted in a 25-μL qPCR reaction volume, which included 12.5 μL of Gene Expression Master Mix (Applied Biosystems), 1.0 μL of 10 μM forward and reverse primers (10 pmol each), 0.5 μL of 5 μM TaqMan probe (2.5 pmol), 5 μL of nuclease-free water, and 5 μL of template cDNA.

qPCR was performed in a Thermal Cycler Dice (Takara Bio Inc., Real Time System III, Kusatsu, Shiga, Japan). The thermal cycling conditions for both assays consisted of pre-heating at 50 °C for 2 min and pre-denaturation at 95 °C for 10 min, followed by 50 cycles of amplification at 95 °C for 15 s, and annealing and extension at 60 °C for 1 min. For each sample, qPCR was performed in technical triplicate for each primer set, and each test was performed twice. Therefore, 6 reactions were obtained for each N1 and N2 assay of each sample.

Every qPCR assay included a negative control (PCR-grade water), and no amplification was observed in either assay. There was no amplification in the negative controls for either the RNA extraction step or the RT step.

As a positive control, a 10-fold serial dilution of double-stranded oligo DNA including both CDC 2019-nCoV_N1 and CDC 2019-nCoV_N2 targets (GeneArt Strings DNA Fragments; Thermo Scientific) was used to generate standard curves (from 10^1^ to 10^4^ copies per 5 μL). The LOQ was 10 copies per reaction with Ct values of 37.7 ± 1.83 for N1 and 38.6 ± 2.28 for N2 primer sets. The theoretical LOQ of the overall method was 4.0 log_10_ copies/L for N1 and N2. The N1 primer set generated a standard curve with *R*^2^ = 0.98 ± 0.04 (*n* = 27 reactions) with an efficiency (mean ± SD) of 106% ± 13.4% (slope = −3.22 ± 0.274; *y* intercept = 40.9% ± 2.01%). The N2 primer set generated a standard curve with *R*^2^ = 0.97 ± 0.05 (*n* = 27 reactions) with an efficiency of 97.7% ± 14.7% (slope = −3.43 ± 0.373; *y* intercept = 42.0% ± 2.54%).

### Counting the positives in repeated qPCR tests

SARS-CoV-2 RNA was detected in many samples in both N1 and N2 qPCR assays, always below the LOQ (see Results and Discussion). Therefore, instead of quantification for SARS-CoV-2 RNA genome copy number, we counted the number of positives in 6 repeated qPCR reactions for each assay. Maximum Ct values were 45 or 46 (Results and Discussion).

### Quality control of the wastewater RT-qPCR assays

PMMoV is the most abundant human fecal RNA virus,^21^ and has recently been used as an internal control for SARS-CoV-2 in wastewater.^7,9-11,20^ When fluorescence reached the threshold during 50 cycles of qPCR in each well of the 96-well plate, we counted it as a positive. We also tested PMMoV RNA by RT-qPCR in the same samples, using 5 μL of template cDNA for all samples, in duplicate, with the primers and probes shown in Table S1 and the same thermal cycling conditions as for SARS-CoV-2 qPCR. We monitored Ct values in each sample to check any loss of SARS-CoV-2 RNA detection.

### Gel electrophoresis and PCR amplicon sequencing

Agarose gel electrophoresis of some samples confirmed the qPCR amplicon sizes: 72 bp for CDC 2019-nCoV_N1 and 67 bp for N2. In addition, qPCR products of N1 and N2 assays of samples 25 and 33 were sequenced by the Molecular Biology Laboratory (Unitech Co., Ltd.; Kashiwa, Chiba, Japan) to confirm whether the qPCR amplified target sites of SARS-CoV-2 RNA. The genome sequence of SARS-CoV-2 Wuhan-Hu-1 strain (GenBank accession No. MN908947.3) was used as a reference.

### COVID-19 Epidemiological data

All patients confirmed positive for COVID-19 by clinical testing in city B were retrospectively interviewed by local public health officers to complete contact tracing. Officers collected recorded symptoms, symptom onset date, contact with other known clinically confirmed cases, and the date of reporting clinical test result (i.e., the date confirmed positive). These data are publicly available. We counted the daily number of cases by the date of symptom onset and by the date reported, from October 2020 to February 2021 (see Results and discussion). For positive asymptomatic cases, the date on which the test result was reported was used as the estimated date of symptom onset.

### Statistical analysis

We investigated whether the SARS-CoV-2 RNA signals in wastewater samples correlated with the epidemiological data. For each wastewater sampling day from October 2020 to February 2021, we compared the number of positives in SARS-CoV-2 RNA PCR reactions and the number of cases who developed symptoms on the same day. We also compared the number of positives in wastewater with the number of cases in clinical testing reported on the same day. Spearman’s rank correlation test was performed in GraphPad Prism 5 software. All statistical tests were two-tailed, with *P* < 0.05 considered as statistically significant.

To evaluate any lag between the SARS-CoV-2 RNA signal in wastewater and the epidemiological data, we performed time-step analyses; we compared the number of positives in wastewater with the number of cases who developed symptoms from 10 days before to 10 days after the wastewater sampling day (i.e., time lag = −10 to +10).

## Results and Discussion

### Concordance between positive counts in qPCR of low-copy-number samples and Poisson distribution

qPCR assays of low-copy-number samples revealed positive reactions in the 12 technical replicates of each ITMN sample (*n* = 12), and positive ratios for each ITMN sample were calculated (Figure 1). Ct values gradually increased as the initial copy number of oligo DNA of SARS-CoV-2 decreased (Figure 1, left). At the same time, the positive ratio decreased. All reactions were positive (*n* = 12, 100%) in all 3 batches when ITMN was ≥6 copies per reaction; around 90% were positive when ITMN was 2 copies; and around 0% to 17% when it was 0.2 copies (Table S2). These positive ratios for each ITMN agree well with those expected from a Poison distribution (Table S3; Figure 1, right). Similar results were reported in previous studies.^15^ From these results, we conclude that by counting the number of positive reactions in repeated qPCR assay, we could trace the dynamics of SARS-CoV-2 RNA in wastewater in the range of 0.1 to 4 copies per reaction of ITMN, which is usually below the LOQ in qPCR and is thus regarded as negative.

**Figure 1.**
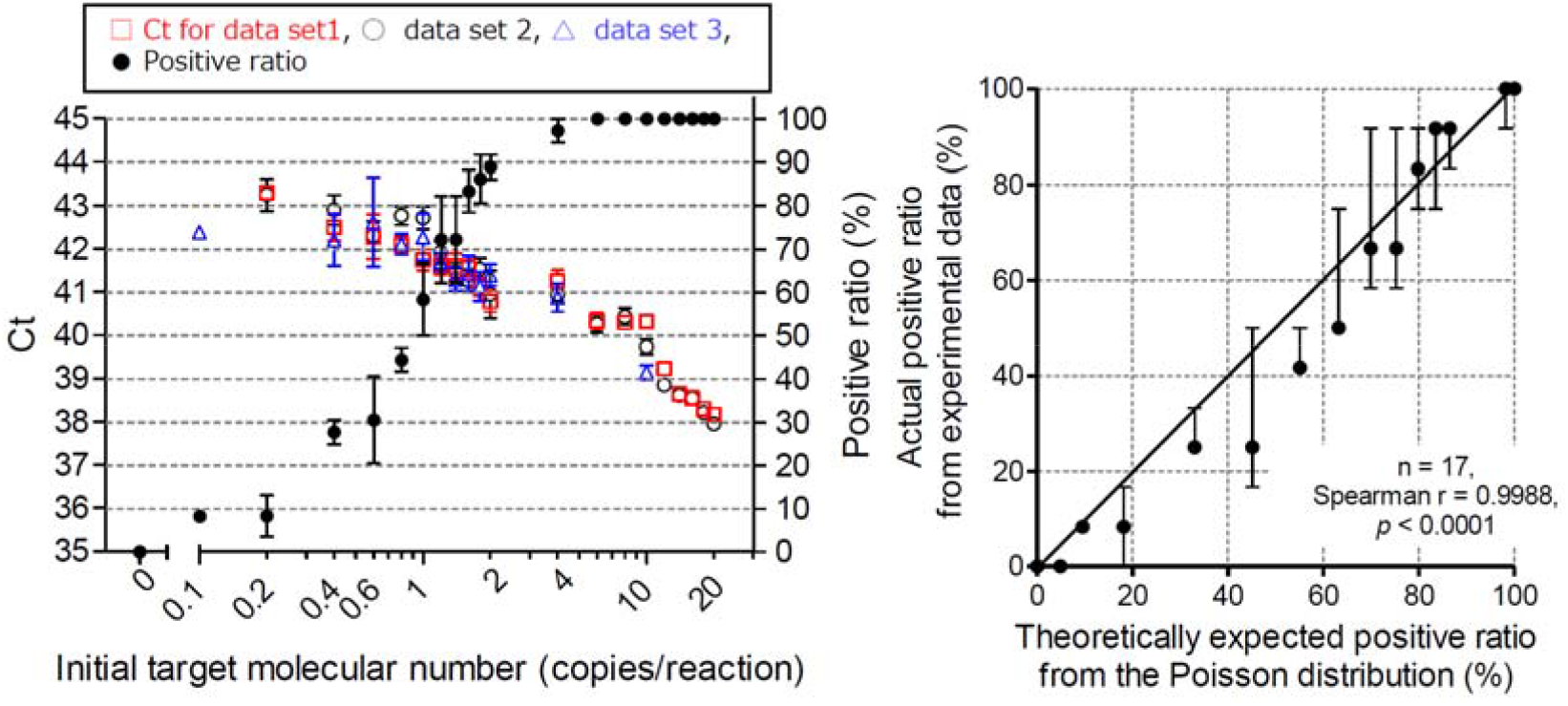
qPCR threshold cycle (Ct) values for low copy numbers of oligo DNA and positive ratios in repeated reactions. N1 assays were performed for oligo DNA, including the target sequence of SARS-CoV-2. Mean ± SEM of the Ct values is shown for each initial target molecular number (ITMN) (0.1–20 copies per reaction) for data sets 1 (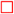, n = 12), 2 (•, n = 12), and 3 (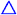, n = 12) (left panel). Mean is calculated for Ct values for which amplification was observed in each batch. Detailed Ct values are shown in SI Table S2. Black plots indicate the average positive ratio of data sets 1, 2, and 3 (Table S3) for each ITMN (n = 36) (right panel).

### Detection of SARS-CoV-2 RNA in wastewater by CDC-N1 and N2 assays

PMMoV was tested as an internal control in all 60 wastewater samples. It was detectable and stable between daily samples, with Ct ranging from 25 to 29 (Figure 2, top; SI Table S4). This result indicates that sample collection, PEG precipitation, RNA extraction, and qPCR processes were not exceptional throughout the sampling campaign.

**Figure 2.**
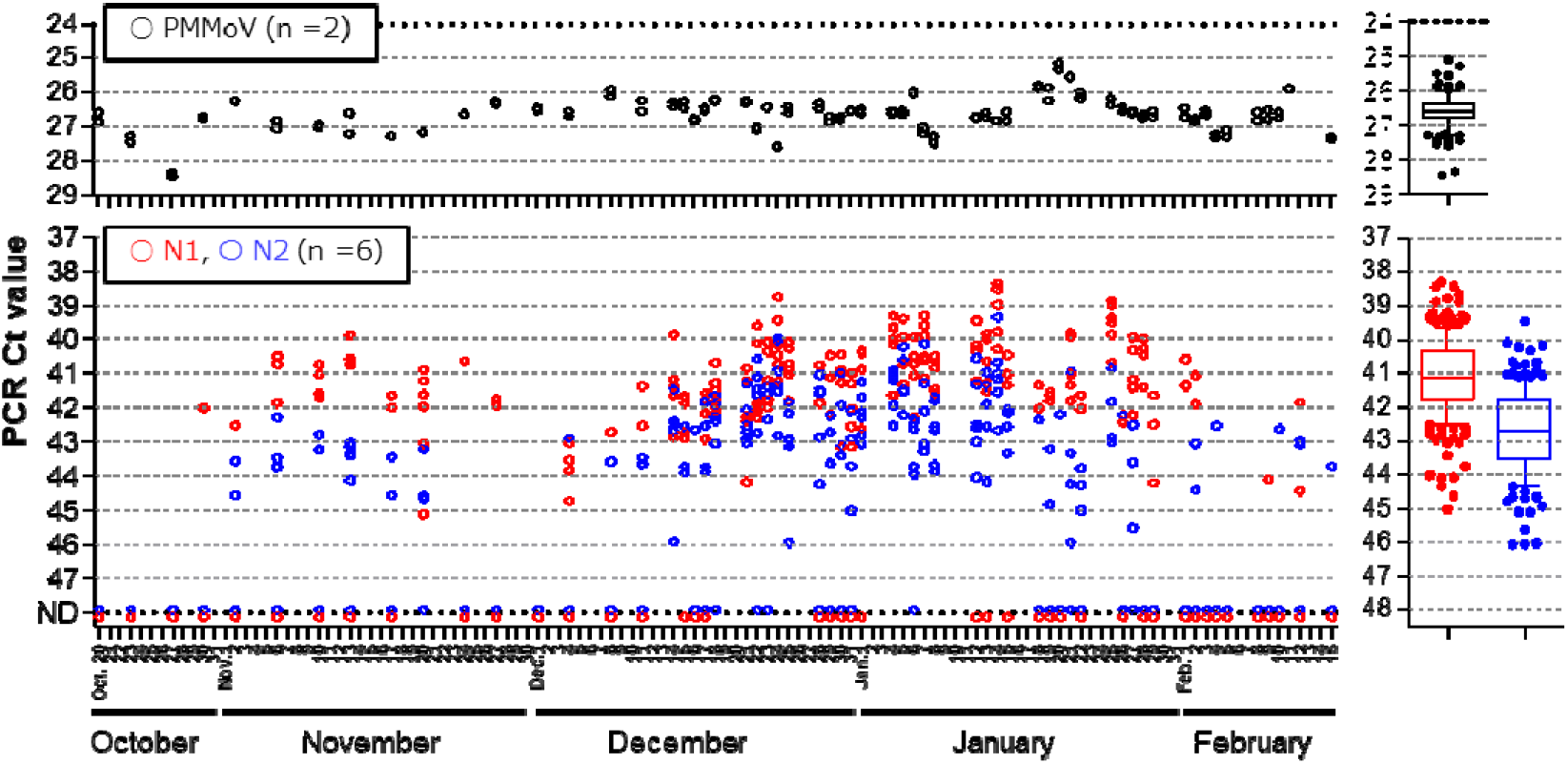
qPCR threshold cycle (Ct) values of PMMoV and SARS-CoV-2 in wastewater samples collected from October 2020 to February 2021. In each sample, PMMoV was tested in duplicate (*n* = 2) and SARS-CoV-2 was tested six times (*n* = 6) with each primer set (CDC-N1 and N2). Each plot indicates the Ct value in each PCR. ND: the sample showed at least 1 negative (*n* = 6). Ct values are shown in SI Table S5.

Maximum Ct values were around 45 in the N1 assay and 46 in the N2 assay (Figure 2, bottom; Table S5). There was no positive detection on the first 3 sample collection days (20, 23, and 27 October). The first positive was detected on 30 October, when the N1 assay (*n* = 6) gave 1 positive and the N2 assay (*n* = 6) gave all negatives.

During early to mid November, both assays gave several positives in repeated PCR reactions on all sampling days, with Ct values of 40 to 45 in the N1 assay and 42 to 45 in the N2 assay (Figure 2, bottom). In late November, the number of positives decreased. But from December, it then increased until mid January 2021, and then again decreased until 15 February. During this period, Ct values were 38 to 45 in the N1 assay and 39 to 46 in the N2 assay. As Ct values of PMMoV were stably detected on all days, fluctuations in the N1 and N2 assays are specific to the SARS-CoV-2 RNA.

Gel electrophoresis confirmed the sizes of the PCR products to be identical to the target sizes (72 bp by N1, 67 bp by N2) by (data not shown). Partial nucleotide sequences of qPCR products from samples 25 and 33 (Table S4) were sequenced and confirmed to be 100% identical to those of the nucleocapsid phosphoprotein of the SARS-CoV-2 genome (GenBank Acc. No. MN908947.3) (SI Figure S2). These results confirm that the target sites of the SARS-CoV-2 genome were amplified by qPCR in this study.

The numbers of positive reactions of the N1 and N2 assays of each wastewater sample are shown in SI Table S5 and Figure 3 (top and middle panels). The numbers of positives for each sample in N1 and N2 assays show a significant positive correlation (SI Figure S3, Spearman *r* = 0.8225; *P* < 0.0001).

**Figure 3.**
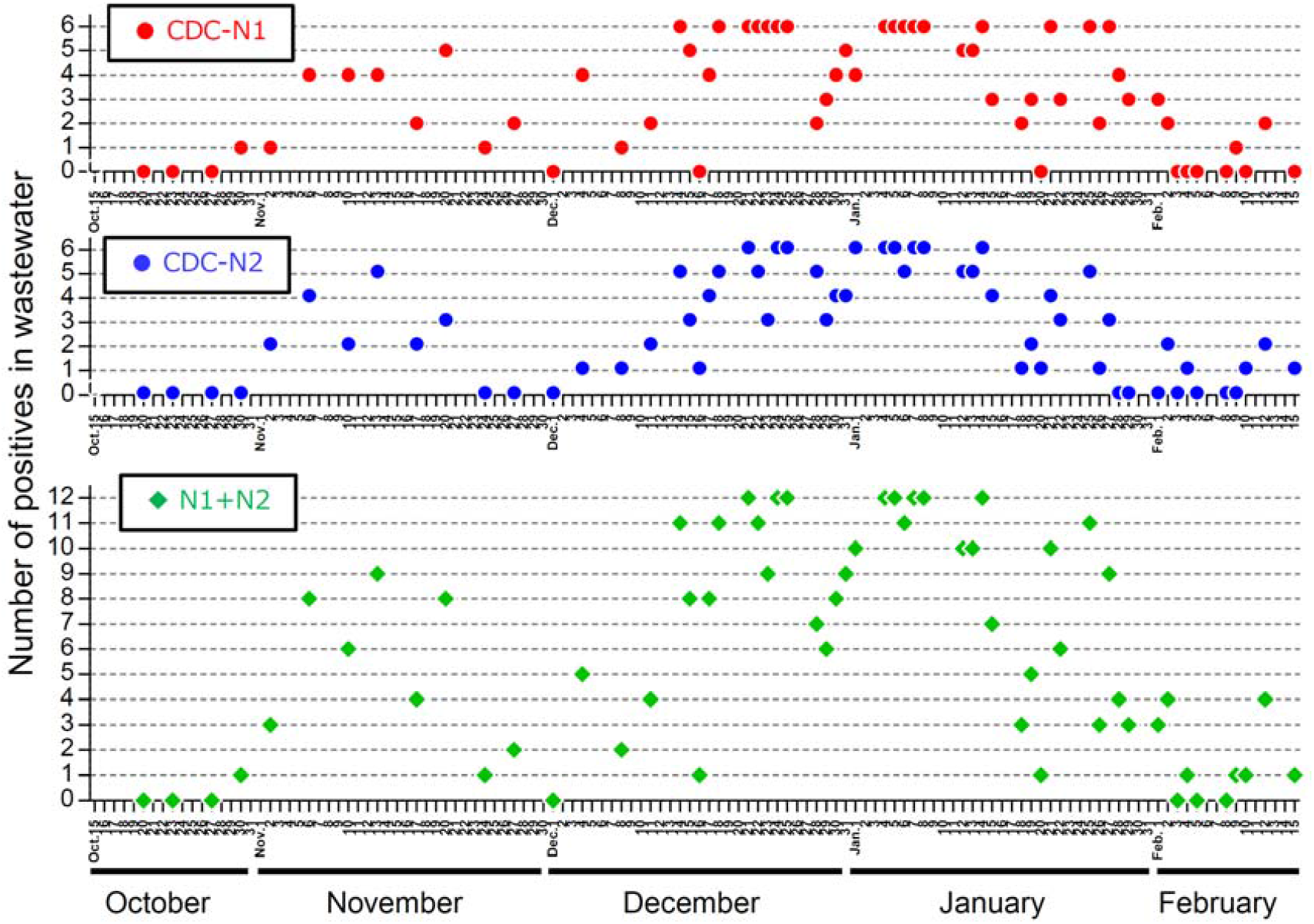
Number of positive reactions by CDC-N1 and N2 assays for SARS-CoV-2 RNA detection in wastewater from WWTP A from October 2020 to February 2021 (*n* = 60). qPCR assay of each sample was performed 6 times with each primer set. Colored plots represent numbers of positive reactions: red, CDC-N1 assay; blue, N2 assay. Green plots: totals in N1 and N2 assays, with a maximum of 12 positives. Positive ratios are also shown.

The sum of the numbers of positive reactions in N1 and N2 assays of each sample fluctuated during the study period (Table S5; Figure 3, bottom panel, N1 + N2). It showed a small peak in November 2020 and a bigger and more persistent peak from December to January (Figure 3), when it sometimes reached the maximum number of positives, i.e., 12. In February, it decreased to as low as 0.

### Correlation between viral signals in wastewater and the number of new cases of COVID-19 in the community

Comparison of viral signals in wastewater with the number of new cases (Table S6) by the date of symptom onset (Figure 4a) and the date reported (Figure 4b) shows clear correlations, with a higher correlation by the former (Fig. 4c: Spearman’s *r* = 0.7538; *P* < 0.0001) than by the latter (Fig. 4d: *r* = 0.5781; *P* < 0.0001). To the best of our knowledge, this is the first study to demonstrate that SARS-CoV-2 RNA in wastewater tracks the daily number of new cases by the date of symptom onset in the community across periods of both increasing and decreasing infection.

**Figure 4.**
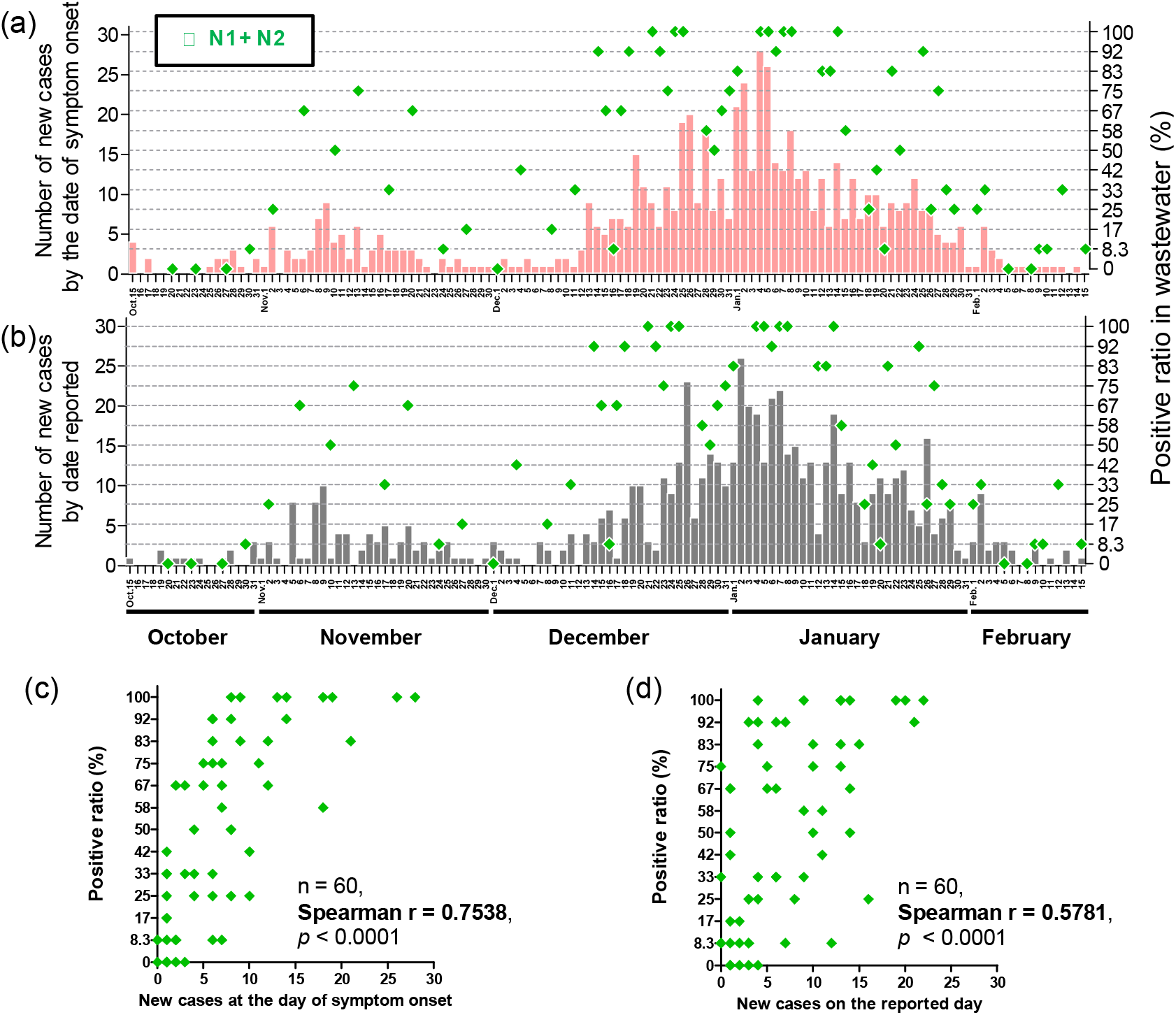
Comparisons between SARS-CoV-2 RNA signal in wastewater and epidemiological data. Comparison between the positive ratio in wastewater (N1 + N2, green plots) and the number of new COVID-19 cases (bars) by (a, c) date of symptom onset and (b, d) date reported. Correlation analysis (Spearman’s *r*) shows a better correlation with (c) the date of symptom onset than (d) with the date reported. Positive ratios in wastewater are taken from Figure 2. Numbers of new cases are detailed in SI Table S6.

### Time-step analysis of correlations between SARS-CoV-2 RNA signal in wastewater and epidemiological data

To evaluate any lag between the SARS-CoV-2 RNA signal in wastewater and the epidemiological data, we performed time-step analyses, comparing the number of positives in wastewater in repeated qPCR reactions on each sampling day with the number of new cases by the date of symptom onset from 10 days before to 10 days after the wastewater sampling day (Figure S4). The number of new cases 2 days before (Figure 5, time lag = −2 days, Spearman’s *r* = 0.7753) and 1 day before (−1, *r* = 0.7279) the sampling day had higher correlations than earlier days (from −10 to −3). And the number of new cases 1 day after (+1, *r* = 0.7177) and 2 days after (+2: *r* = 0.7283) the sampling day had higher correlations than later days (from +3 to +10, *r* < 0.7). These results indicate that COVID-19 patients excreted more virus in their feces in the period from 2 days before to 2 days after symptom onset than on other days, which wastewater surveillance could detect.

**Figure 5.**
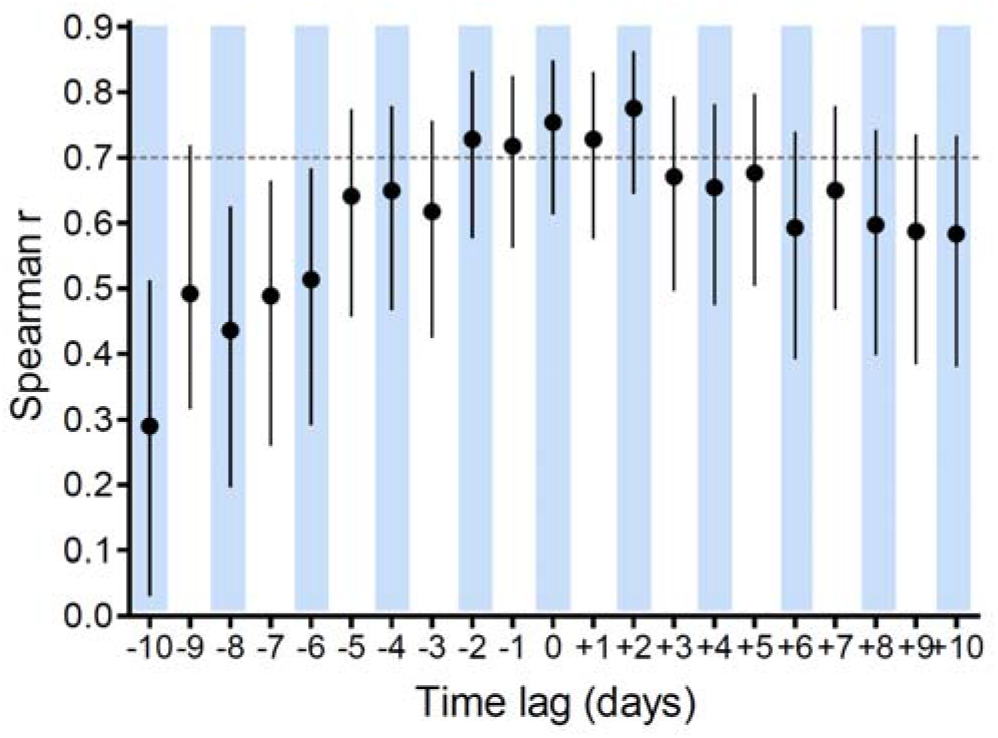
Time-step analysis of correlation between SARS-CoV-2 RNA signal in wastewater and epidemiological data. Viral RNA signals in wastewater are compared with numbers of new COVID-19 cases by the date of symptom onset from 10 days before (−10) to 10 days after (+10) the wastewater sampling day (0). Spearman’s *r* values are plotted with 95% confidence interval.

Previous studies have reported that SARS-CoV-2 RNA concentrations in wastewater were correlated with epidemiological data such as the number of clinically confirmed cases by the date of specimen collection or of test result reporting,^4-8,10^ but did not investigate the correlation with the number of cases by the date of symptom onset. So far, only one study has investigated this correlation, and reported that wastewater detection of the SARS-CoV-2 RNA trailed symptom onset by 5 days.^5^ Here, in contrast, the number of positives in repeated PCR reactions in wastewater was significantly correlated with the new cases at the date of symptom onset within 2 days before to 2 days after. The difference in results might be explained by the fact that we started sampling before the third wave of the pandemic began, and captured the initial phase of the wave, while Nemudryi et al. (2020) started sampling after the peak of the infection, so they missed the chance to detect the viral RNA signal in wastewater in the initial increasing phase of infection.^5^

### How much faster can WBE track COVID-19 in the community than clinical testing?

How much sooner did the SARS-CoV-2 RNA concentration in wastewater than the date of result reporting or specimen collection in the public health system? On average, a person develops symptoms 4 to 5 days after infection.^22^ Our results indicate that SARS-CoV-2 RNA concentrations in feces are highest during 2 days either side of symptom onset (Figure 6). In regions where the time lag between the date of symptom onset and the date of clinical test result reporting is large, WBE, if analyzed and reported on the same day as sampling, will give the earliest estimate of viral spread in the community. This is why WBE has an advantage over clinical testing in the public health system.

**Figure 6.**
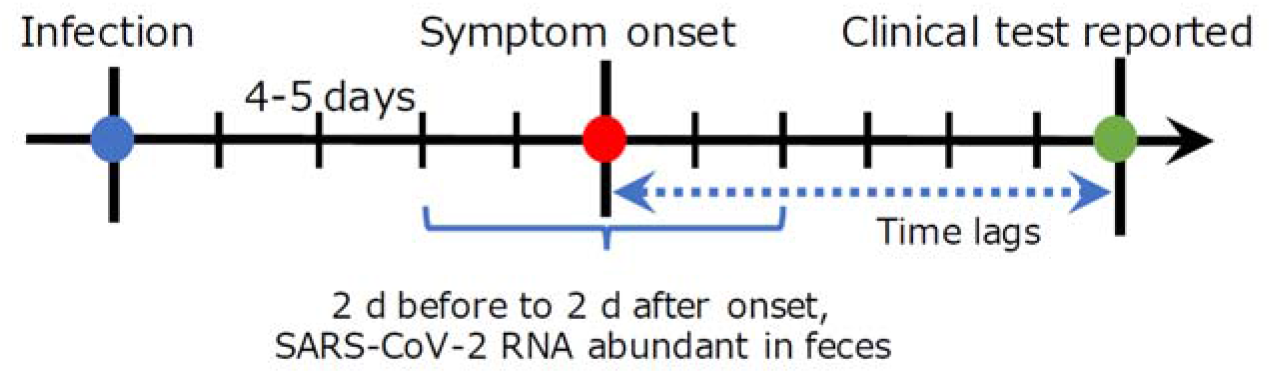
Timeline of clinical testing and wastewater surveillance.

### Sensitivity of the positive count for SARS-CoV-2 RNA detection in wastewater

Throughout the sampling period, N1 and N2 assays in wastewater samples showed significant correlation with the number of new cases in the range of 0 to 30 (Figure 4). The population of city B is about 300,000, and WWTP A serves about one-third of it. This means that SARS-CoV-2 RNA in wastewater could track the infection dynamics in the community even when the number of new cases ranges from 0 to 10 per 100,000 population. However, the possibility of asymptomatic patients means that the actual number of cases might be larger.

### Implications of viral shedding in feces

These results indicate that COVID-19 patients excrete more virus in their feces during 2 days either side of symptom onset than earlier or later (Figure 5). Clinical studies have reported virus concentrations in feces several days after symptom onset^23^ or after admission to hospital^24,25^. Under the assumption that virus shedding in feces starts from the time of symptom onset, re-analysis of patient data using a shedding dynamics model indicated that the SARS-CoV-2 RNA concentration in feces increases rapidly after symptom onset.^26^ However, no clinical data on viral shedding in feces from infection to symptom onset are reported. Still, it is possible that the peak occurs before symptom onset. To understand what the viral RNA signal in wastewater represents, the peak timing of viral shedding in feces is essential information. More fecal shedding data are needed.

As patients likely shed SARS-CoV-2 in feces for weeks,^24,26,27^ it is somewhat surprising that SARS-CoV-2 RNA signals in wastewater significantly correlated with daily confirmed new cases in this study. But these results indicate that the magnitude of fecal shedding might be substantially higher around the day of symptom onset. Another possibility is that the virus RNA in wastewater is derived not only from feces, but also from the respiratory system and saliva: virus excreted during brushing the teeth, gargling, and rinsing could mix with virus in feces and travel to the WWTP. Viral concentrations in throat swabs from patients were highest immediately after onset, and therefore it is estimated that infectiousness is highest from 2 to 1 day before the onset date.^28^ The period of highest infectiousness might also be the period of highly detectable virus RNA in wastewater.

### Usefulness and limitation of the positive count method

Although SARS-CoV-2 RNA concentrations in wastewater were lower than the LOQ, the number of positives in repeated RT-qPCR reactions could trace the prevalence of COVID-19 in the community. In conventional qPCR testing, the limit of quantification is usually 5 to 10 copies per reaction. In contrast, the positive count method could distinguish lower copy numbers, i.e., 0.1 to 4 copies per reaction, which means that it increases the sensitivity of qPCR by 50 to 100 times to reveal the trend of viral RNA concentrations in wastewater. For the purpose of tracking the virus infection dynamics in the community, exact values of virus concentrations are not always necessary; it is enough to see the change of viral RNA levels in wastewater, for which the positive count method is useful.

We used the positive count method with RT-qPCR of the supernatant fraction of wastewater. This method can also be used with the solid fraction of wastewater. Recent studies have improved the sensitivity of viral RNA detection in wastewater by RT-qPCR by concentrating virus from the solid fraction of raw influent or primary sludge rather than the water fraction.^6,8-11^ However, in regions where the virus concentration in wastewater is not high enough for quantification from the solid fraction, the positive count methods might be useful to reveal changes in viral RNA levels in wastewater.

The idea of the positive count method is the same as that for the quantification of droplet digital PCR. Therefore, we expect to see use of the droplet digital PCR in future studies to show the trend of SARS-CoV-2 RNA at low copy numbers in wastewater, as the positive count method has done here.

## Data Availability

All data produced in the present work are contained in the manuscript

## Acknowledgments

This work was supported by grants from the GAP Fund Program of Kyoto University; the Keihanshin Consortium for Fostering the Next Generation of Global Leaders in Research (K-CONNEX), established by Human Resource Development Program for Science and Technology, MEXT; JST-Mirai Program Grant Number JPMJMI18DA, from the Japan Science and Technology Agency; and JST Adaptable and Seamless Technology transfer Program through Target-driven R&D (A-STEP) Grant Number JPMJTM20Y5, from the Japan Science and Technology Agency. We thank all staff of local wastewater bureaus for providing meteorological data, helping with the sampling, and providing water quality and quantity data. We also thank Dr. Akihiko Hata for technical support on pretreatment of samples.

