## Supporting information for "Tracking community infection dynamics of COVID-19 by monitoring SARS-CoV-2 RNA in wastewater, counting positive reactions by qPCR"

**CONTENTS**

**24 pages.**

Page 3. Table S1. Details of primers and probes of qPCR assays

Page 4. Table S2. Positive number and ratio in qPCR for low-target copy number samples

Page 7. Table S3. Analysis of the coherence to Poisson distribution of various CDC-N1 qPCR assays containing low initial copy number of target molecule

Page 8. Table S4. Ct value of PMMoV RNA in wastewater of WWTP A

Page 10. Table S5. Details of detection of SARS-CoV-2 RNA in wastewater of WWTP A

Page 13. Table S6. Comparison between epidemiological data of COVID-19 cases and wastewater-based epidemiology data

Page 20. Figure S1. Poisson distribution of the probability for the number of molecules per reaction in qPCR

Page 21. Figure S2. Sequence of qPCR products from wastewater samples

Page 22. Figure S3. Correlation on positive numbers between CDC-N1 and CDC-N2 assays for wastewater of WWTP A

Page 23. Figure S4. Time-step analysis for the correlation between new cases in clinical test and positive numbers of N1+N2 assay in wastewater

Page 24. References

**Table S1.** Details of primers and probes of qPCR assays.

| Assay | Function | Name | Sequence (5′–3′) | Product length (bp) |
| --- | --- | --- | --- | --- |
| SARS-CoV-2  (CDC-N1 and CDC-N2)^a^ | Forward primer | 2019-nCoV_N1-F | GACCCCAAAATCAGCGAAAT | 72 |
|  | Reverse primer | 2019-nCoV_N1-R | TCTGGTTACTGCCAGTTGAATCTG |  |
|  | TaqMan probe | 2019-nCoV_N1-P | FAM-ACCCCGCATTACGTTTGGTGGACC-BHQ1 |  |
|  | Forward | 2019-nCoV_N2-F | TTACAAACATTGGCCGCAAA | 67 |
|  | Reverse | 2019-nCoV_N2-R | GCGCGACATTCCGAAGAA |  |
|  | Probe | 2019-nCoV_N2-P | FAM-ACAATTTGCCCCCAGCGCTTCAG-BHQ1 |  |
| PMMoV^b^ | Forward | PMMoV-FP1 | GAGTGGTTTGACCTTAACGTTTGA |  |
|  | Reverse | PMMoV-RP1 | TTGTCGGTTGCAATGCAAGT | 68 |
|  | TaqMan MGB probe | PMMoV-Probe1 | FAM-CCTACCGAAGCAAATG-MGB-NFQ |  |

PMMoV: Pepper Mild Mottle Virus

^a^ Reference 1

^b^ Reference 2 and 3

**Table S2.** Positive number and ratio by qPCR of low-target-copy-number samples.

| Data set 1 | Ct value for N1 (*n* = 12) | | | | | | | | | | | |  | |
| --- | --- | --- | --- | --- | --- | --- | --- | --- | --- | --- | --- | --- | --- | --- |
| Initial copy number per reaction  (copies/5 μL) | 1 | 2 | 3 | 4 | 5 | 6 | 7 | 8 | 9 | 10 | 11 | 12 | Positives number (*n* = 12) | Positive ratio  (%) |
| 20 | 37.83 | 38.07 | 37.3 | 37.96 | 38 | 38.29 | 37.75 | 37.99 | 37.8 | 38.32 | 38.07 | 38.3 | 12 | 100 |
| 18 | 38.37 | 38.28 | 38.06 | 38.16 | 38.11 | 38.46 | 37.98 | 37.73 | 38.4 | 38.06 | 38.19 | 39 | 12 | 100 |
| 16 | 38.79 | 38.64 | 38.02 | 38.65 | 38.4 | 37.98 | 38.93 | 38.02 | 38.27 | 38.39 | 38.64 | 38.72 | 12 | 100 |
| 14 | 38.03 | 39.02 | 37.78 | 38.59 | 38.61 | 38.8 | 39 | 38.9 | 38.75 | 39.37 | 38.53 | 38.62 | 12 | 100 |
| 12 | 38.81 | 38.83 | 39.67 | 39.33 | 39.12 | 38.47 | 38.45 | 38.76 | 39.27 | 38.55 | 38.76 | 39.4 | 12 | 100 |
| 10 | 40.13 | 39.58 | 39.56 | 39.51 | 39.92 | 38.59 | 39.79 | 40.02 | 40.5 | 39.57 | 39.48 | 39.12 | 12 | 100 |
| 8 | 41.5 | 40.5 | 40.07 | 40.75 | 39.32 | 39.15 | 39.67 | 41.15 | 41.42 | 40.23 | 40.6 | 41.78 | 12 | 100 |
| 6 | 40.03 | 41.14 | 40.15 | 39.14 | 39.59 | 39.66 | 41.02 | 40.1 | 39.53 | 39.99 | 39.86 | 40.14 | 12 | 100 |
| 4 | 41.45 | 40.87 | 40.36 | 42.02 | 40.67 | 41.1 | 40.4 | 41.18 | 41.14 | 42.33 | 40.56 | 40.43 | 12 | 100 |
| 2 | 40.89 | – | 41.26 | 41.88 | 41.13 | 39.65 | 36.06 | 40.36 | 40.88 | 46.99 | 41.45 | 40.72 | 11 | 91.7 |
| 1.8 | 42.38 | 41.97 | 42.04 | 42.25 | 40.17 | 40.7 | 42.39 | 41.08 | 40.57 | – | 43.14 | 42.6 | 11 | 91.7 |
| 1.6 | 39.74 | 40.85 | – | – | 40.97 | 42.26 | 41.09 | 40.71 | 40.56 | 41.86 | 41.13 | 42.33 | 10 | 83.3 |
| 1.4 | 42.31 | 40.76 | 40.49 | 41.23 | 41.97 | 42.58 | 41.97 | 41.17 | 41.17 | 40.96 | 41.31 | – | 11 | 91.7 |
| 1.2 | 42.19 | 41.2 | – | 42.2 | 42.99 | – | – | – | 42.3 | 42.24 | 39.86 | 42.08 | 8 | 66.7 |
| 1 | 42.99 | 42.72 | 44.26 | – | 44.12 | 41.52 | – | 41.64 | 41.86 | 41.86 | – | 42.26 | 9 | 75.0 |
| 0.8 | – | 42.8 | 42.91 | – | – | – | 42.2 | – | – | 42.85 | 42.12 | 42.65 | 6 | 50.0 |
| 0.6 | – | – | – | 40.87 | 41.3 | – | 44.31 | 41.64 | 42.79 | 42.78 | – | – | 6 | 50.0 |
| 0.4 | – | 42.89 | – | 42.42 | – | – | – | 42.63 | – | – | – | – | 3 | 25.0 |
| 0.2 | – | – | – | – | – | – | – | – | – | – | – | 42.8 | 1 | 8.3 |
| 0 (without DNA) | – | – | – | – | – | – | – | – | – | – | – | – | 0 | 0.0 |

"–": not detected (Ct > 50)

**Table S2 (Continued).** Positive number and ratio by qPCR of low-target-copy-number samples.

| Data set 2 | Ct value for N1 (*n* = 12) | | | | | | | | | | | |  |  |
| --- | --- | --- | --- | --- | --- | --- | --- | --- | --- | --- | --- | --- | --- | --- |
| Initial copy number per reaction  (copies/5 μL) | 1 | 2 | 3 | 4 | 5 | 6 | 7 | 8 | 9 | 10 | 11 | 12 | Positives number (*n* = 12) | Positive ratio  (%) |
| 20 | 37.69 | 38.06 | 37.93 | 38.26 | 37.59 | 38.24 | 38.24 | 37.87 | 38.77 | 38.58 | 38.65 | 37.63 | 12 | 100 |
| 18 | 39 | 38.74 | 37.97 | 37.94 | 38.25 | 38.69 | 37.79 | 38.44 | 37.85 | 38.74 | 38.11 | 38.23 | 12 | 100 |
| 16 | 38.56 | 37.96 | 38.73 | 38.26 | 39.49 | 38.79 | 38.38 | 38.01 | 38.37 | 38.35 | 38.16 | 38.29 | 12 | 100 |
| 14 | 38.87 | 39.32 | 38.19 | 39.37 | 38.1 | 38.7 | 37.95 | 39.25 | 38.85 | 38.55 | 38.07 | 37.93 | 12 | 100 |
| 12 | 39.48 | 39.22 | 39.08 | 38.62 | 39.23 | 39.75 | 39.21 | 39.25 | 38.27 | 39.46 | 39.63 | 40.04 | 12 | 100 |
| 10 | 39.28 | 41.25 | 40.65 | 39.79 | 39.8 | 40.62 | 40.89 | 39.74 | 39.98 | 40.79 | 40.11 | 40.27 | 12 | 100 |
| 8 | 40.47 | 40 | 40.76 | 40.92 | 40.16 | 39.9 | 39.86 | 40.78 | 39.86 | 40.7 | 40.36 | 40.58 | 12 | 100 |
| 6 | 40.55 | 40.6 | 39.42 | 39.57 | 39.28 | 40.16 | 41.92 | 39.22 | 39.96 | 40.3 | 41.2 | 40.67 | 12 | 100 |
| 4 | 40.09 | 41.9 | 40.64 | 41.58 | 40.1 | 41.32 | 40 | 42.75 | 40.01 | 40.99 | 40.89 | 41.42 | 12 | 100 |
| 2 | 39.56 | 40.5 | 39.45 | 41.7 | 40.63 | 40.74 | – | 40.74 | 40.51 | 41.86 | 39.49 | 42.28 | 11 | 91.7 |
| 1.8 | 41.12 | 40.59 | 40.79 | 40.2 | – | 40.22 | 41.08 | 39.66 | 42.02 | 41.8 | 41.01 | 41.65 | 11 | 91.7 |
| 1.6 | 42.05 | 39.92 | 41.86 | 41.78 | 40.72 | – | 40.79 | 42.27 | 41.4 | 41.88 | 41.9 | 42.77 | 11 | 91.7 |
| 1.4 | – | 42.14 | 41.14 | – | 40.54 | – | 41.01 | – | 41.72 | 40.17 | 41.43 | – | 7 | 58.3 |
| 1.2 | 42.14 | 40.66 | 41.12 | 41.41 | 41.01 | 41.09 | 41.07 | 42.47 | 41.62 | – | 42.14 | 41.12 | 11 | 91.7 |
| 1 | 43.22 | – | 41.67 | 40.9 | 41.85 | – | – | – | – | – | 41.07 | 42.05 | 6 | 50.0 |
| 0.8 | – | 42.34 | – | 42.48 | – | 41.84 | – | – | – | 41.61 | – | 43.16 | 5 | 41.7 |
| 0.6 | – | – | 41.7 | 41.82 | – | – | – | – | – | – | 43.31 | – | 3 | 25.0 |
| 0.4 | – | – | 42.11 | 42.73 | 42.29 | – | – | – | – | – | 43.59 | – | 4 | 33.3 |
| 0.2 | 43.32 | – | – | – | – | – | – | – | 43.33 | – | – | – | 2 | 16.7 |
| 0 (without DNA) | – | – | – | – | – | – | – | – | – | – | – | – | 0 | 0.0 |

"–": not detected (Ct > 50)

**Table S2 (Continued).** Positive number and ratio by qPCR of low-target-copy-number samples.

| Data set 3 | Ct value for N1 (*n* = 12) | | | | | | | | | | | |  |  |
| --- | --- | --- | --- | --- | --- | --- | --- | --- | --- | --- | --- | --- | --- | --- |
| Initial copy number per reaction  (copies/5 μL) | 1 | 2 | 3 | 4 | 5 | 6 | 7 | 8 | 9 | 10 | 11 | 12 | Positives number (*n* = 12) | Positive ratio  (%) |
| 10 | 39.88 | 38.4 | 39.13 | 39.56 | 38.94 | 38.16 | 39.65 | 38.47 | 39.64 | 39.76 | 39.03 | 39.19 | 12 | 100 |
| 4 | 42.22 | 39.9 | 41.92 | 40.28 | 40.83 | 39.69 | 40.83 | – | 39.85 | 40.18 | 40.9 | 42.94 | 11 | 91.7 |
| 2 | – | 40.9 | 41.72 | 42.48 | 42.08 | 41.61 | 40.92 | 40.56 | – | 41.67 | 41.98 | 39.97 | 10 | 83.3 |
| 1.8 | 40.42 | – | 42.76 | – | 40.76 | 39.32 | – | 41.72 | 40.62 | 41.91 | 41.73 | 40.99 | 9 | 75.0 |
| 1.6 | 39.36 | – | – | 42.44 | 41.03 | 40.85 | 42.43 | 39.71 | 42.75 | 42.15 | – | 42.1 | 9 | 75.0 |
| 1.4 | 39.74 | 41.58 | – | – | 40.94 | 40.86 | 41.95 | 41.73 | 41.62 | – | – | 41.85 | 8 | 66.7 |
| 1.2 | 41.94 | 41.14 | 42.1 | – | – | – | – | 41.67 | 41.85 | 40.92 | – | 42.31 | 7 | 58.3 |
| 1 | – | – | 40.89 | 41.45 | 44.36 | – | 43.41 | 41.9 | – | – | – | 41.61 | 6 | 50.0 |
| 0.8 | – | 41.72 | – | – | 41.58 | – | – | – | 42.47 | 41.94 | – | 42.85 | 5 | 41.7 |
| 0.6 | – | – | – | – | – | 43.65 | – | 41.59 | – | – | – | – | 2 | 16.7 |
| 0.4 | – | 41.27 | 42.03 | – | – | 43.32 | – | – | – | – | – | – | 3 | 25.0 |
| 0.2 | – | – | – | – | – | – | – | – | – | – | – | – | 0 | 0.0 |
| 0.1 | – | – | – | – | – | 42.39 | – | – | – | – | – | – | 1 | 8.3 |
| 0.01 | – | – | – | – | – | – | – | – | – | – | – | – | 0 | 0.0 |
| 0 (without DNA) | – | – | – | – | – | – | – | – | – | – | – | – | 0 | 0.0 |

"–": not detected (Ct > 50)

**Table S3. Analysis of agreement with Poisson distribution of various CDC-N1 qPCR assays containing low initial copy number of target molecule.**

|  | Expected initial target molecular number (ITMN) per reaction (copies/5 μL) *^a^* | | | | | | | | | | | | | | |
| --- | --- | --- | --- | --- | --- | --- | --- | --- | --- | --- | --- | --- | --- | --- | --- |
|  | 0 | 0.01 | 0.1 | 0.2 | 0.4 | 0.6 | 0.8 | 1 | 1.2 | 1.4 | 1.6 | 1.8 | 2 | 4 | 10 |
| P(X≥1) *^b^* |  | 0.00995 | 0.09516 | 0.18127 | 0.32968 | 0.45119 | 0.55067 | 0.63212 | 0.69881 | 0.75340 | 0.79810 | 0.83470 | 0.86466 | 0.98168 | 0.99995 |
| P(X=0) *^b^* |  | 0.99005 | 0.90484 | 0.81873 | 0.67032 | 0.54881 | 0.44933 | 0.36788 | 0.30119 | 0.24660 | 0.20190 | 0.16530 | 0.13534 | 0.01832 | 0.00005 |
| Expected positive counts *^b^* | 0 | 0.1 | 1.1 | 2.2 | 4.0 | 5.4 | 6.6 | 7.6 | 8.4 | 9.0 | 9.6 | 10.0 | 10.4 | 11.8 | 12.0 |
| Expected negative counts *^b^* |  | 11.9 | 10.9 | 9.8 | 8.0 | 6.6 | 5.4 | 4.4 | 3.6 | 3.0 | 2.4 | 2.0 | 1.6 | 0.2 | 0.0 |
| Expected positive ratio (%) |  | **1.0** | **9.5** | **18.1** | **33.0** | **45.1** | **55.1** | **63.2** | **69.9** | **75.3** | **79.8** | **83.5** | **86.5** | **98.2** | **100** |
| Actual positive ratio (%) *^c^* (data set 1) | 0.0 |  |  | 8.3 | 25.0 | 50.0 | 50.0 | 75.0 | 66.7 | 91.7 | 83.3 | 91.7 | 91.7 | 100 | 100 |
| (data set 2) | 0.0 |  |  | 16.7 | 33.3 | 25.0 | 41.7 | 50.0 | 91.7 | 58.3 | 91.7 | 91.7 | 91.7 | 100 | 100 |
| (data set 3) | 0.0 | 0.0 | 8.3 | 0.0 | 25.0 | 16.7 | 41.7 | 50.0 | 58.3 | 66.7 | 75.0 | 75.0 | 83.3 | 91.7 | 100 |
| Average | 0.0 | **0.0** | **8.3** | **8.3** | **27.8** | **30.6** | **44.5** | **58.3** | **72.7** | **72.7** | **83.3** | **86.1** | **88.9** | **97.2** | **100** |

a: Concentration of oligo DNA including N1 qPCR target sequence.

b: Theoretically expected values from the probability of the Poisson distribution and sample number (n = 12) for each ITMN. Positive and negative counts are calculated from P(x = 0) and P(x ≥ 1), respectively, as shown in Figure S1.

c: Values derived from experimental data (n = 12) (Table S2).

**Table S4.** Ct value of PMMoV RNA in wastewater of WWTP A.

| Date | Sample ID | Threshold cycle (Ct) value (*n* = 2) | |
| --- | --- | --- | --- |
|  |  | 1 | 2 |
| 20201020 | 1 | 26.85 | 26.57 |
| 20201023 | 2 | 27.44 | 27.24 |
| 20201027 | 3 | 28.46 | 28.35 |
| 20201030 | 4 | 26.75 | 26.68 |
| 20201102 | 5 | 26.22 | 26.23 |
| 20201106 | 6 | 27.02 | 26.81 |
| 20201110 | 7 | 26.98 | 26.92 |
| 20201113 | 8 | 26.60 | 27.18 |
| 20201117 | 9 | 27.25 | 27.25 |
| 20201120 | 10 | 27.14 | 27.14 |
| 20201124 | 11 | 26.62 | 26.61 |
| 20201127 | 12 | 26.33 | 26.25 |
| 20201201 | 13 | 26.50 | 26.42 |
| 20201204 | 14 | 26.67 | 26.55 |
| 20201208 | 15 | 25.92 | 26.08 |
| 20201211 | 16 | 26.21 | 26.53 |
| 20201214 | 17 | 26.26 | 26.38 |
| 20201215 | 18 | 26.24 | 26.43 |
| 20201216 | 19 | 26.75 | 26.81 |
| 20201217 | 20 | 26.43 | 26.51 |
| 20201218 | 21 | 26.18 | 26.22 |
| 20201221 | 22 | 26.29 | 26.22 |
| 20201222 | 23 | 27.00 | 27.06 |
| 20201223 | 24 | 26.41 | 26.41 |
| 20201224 | 25 | 27.58 | 27.56 |
| 20201225 | 26 | 26.40 | 26.57 |
| 20201228 | 27 | 26.44 | 26.30 |
| 20201229 | 28 | 26.67 | 26.85 |
| 20201230 | 29 | 26.78 | 26.67 |
| 20201231 | 30 | 26.50 | 26.53 |
| 20210101 | 31 | 26.60 | 26.47 |
| 20210104 | 32 | 26.54 | 26.61 |
| 20210105 | 33 | 26.53 | 26.63 |
| 20210106 | 34 | 26.00 | 25.96 |
| 20210107 | 35 | 27.16 | 27.01 |
| 20210108 | 36 | 27.26 | 27.47 |
| 20210112 | 37 | 26.68 | 26.73 |
| 20210113 | 38 | 26.72 | 26.58 |
| 20210114 | 39 | 26.81 | 26.81 |
| 20210115 | 40 | 26.55 | 26.82 |
| 20210118 | 41 | 25.86 | 25.78 |
| 20210119 | 42 | 25.85 | 26.21 |

**Table S4 (Continued).** Ct value of PMMoV RNA in wastewater of WWTP A.

| Date | Sample ID | Ct value (*n* = 2) | |
| --- | --- | --- | --- |
|  |  | 1 | 2 |
| 20210120 | 43 | 25.11 | 25.30 |
| 20210121 | 44 | 25.55 | 25.50 |
| 20210122 | 45 | 26.13 | 26.00 |
| 20210125 | 46 | 26.16 | 26.33 |
| 20210126 | 47 | 26.40 | 26.52 |
| 20210127 | 48 | 26.53 | 26.60 |
| 20210128 | 49 | 26.63 | 26.72 |
| 20210129 | 50 | 26.69 | 26.54 |
| 20210201 | 51 | 26.70 | 26.44 |
| 20210202 | 52 | 26.74 | 26.85 |
| 20210203 | 53 | 26.64 | 26.52 |
| 20210204 | 54 | 27.19 | 27.31 |
| 20210205 | 55 | 27.28 | 27.09 |
| 20210208 | 56 | 26.56 | 26.81 |
| 20210209 | 57 | 26.49 | 26.78 |
| 20210210 | 58 | 26.57 | 26.71 |
| 20210212 | 59 | 25.89 | 25.87 |
| 20210215 | 60 | 27.36 | 27.28 |

**Table S5.** Details of detection of SARS-CoV-2 RNA in wastewater of WWTP A.

| Date | Sample ID | Threshold cycle (Ct) value for N1 (*n* = 6) | | | | | | Positive number  for N1 | Ct value for N2 (*n* = 6) | | | | | | Positive number  for N2 |
| --- | --- | --- | --- | --- | --- | --- | --- | --- | --- | --- | --- | --- | --- | --- | --- |
|  |  | 1 | 2 | 3 | 4 | 5 | 6 |  | 1 | 2 | 3 | 4 | 5 | 6 |  |
| 20201020 | 1 | – | – | – | – | – | – | 0 | – | – | – | – | – | – | 0 |
| 20201023 | 2 | – | – | – | – | – | – | 0 | – | – | – | – | – | – | 0 |
| 20201027 | 3 | – | – | – | – | – | – | 0 | – | – | – | – | – | – | 0 |
| 20201030 | 4 | – | – | – | – | – | 41.90 | 1 | – | – | – | – | – | – | 0 |
| 20201102 | 5 | – | – | – | – | – | 42.41 | 1 | – | – | 44.64 | 43.65 | – | – | 2 |
| 20201106 | 6 | 40.41 | – | – | 41.75 | 40.62 | 40.38 | 4 | 43.83 | – | – | 43.56 | 42.38 | 43.8 | 4 |
| 20201110 | 7 | 41.48 | – | 40.92 | 40.63 | – | 41.59 | 4 | – | – | 43.31 | – | – | 42.88 | 2 |
| 20201113 | 8 | – | 39.78 | 40.48 | 40.59 | – | 40.65 | 4 | 43.21 | 43.12 | 43.50 | 44.22 | 43.44 | – | 5 |
| 20201117 | 9 | – | 41.53 | – | 41.89 | – | – | 2 | 44.64 | – | – | – | – | 43.53 | 2 |
| 20201120 | 10 | 42.95 | 41.51 | 40.79 | – | 41.86 | 41.1 | 5 | – | 44.67 | – | 44.76 | 43.29 | – | 3 |
| 20201124 | 11 | – | – | 40.52 | – | – | – | 1 | – | – | – | – | – | – | 0 |
| 20201127 | 12 | – | – | 41.64 | – | 41.83 | – | 2 | – | – | – | – | – | – | 0 |
| 20201201 | 13 | – | – | – | – | – | – | 0 | – | – | – | – | – | – | 0 |
| 20201204 | 14 | 43.73 | – | 44.61 | – | 42.91 | 43.42 | 4 | – | – | – | – | 43.00 | – | 1 |
| 20201208 | 15 | – | – | – | – | 42.6 | – | 1 | – | – | – | – | 43.67 | – | 1 |
| 20201211 | 16 | – | – | 41.27 | – | – | 42.42 | 2 | – | – | – | – | 43.56 | 43.74 | 2 |
| 20201214 | 17 | 39.76 | 41.28 | 41.07 | 42.76 | 42.59 | 41.53 | 6 | 42.48 | 46.00 | – | 42.78 | 42.58 | 41.57 | 5 |
| 20201215 | 18 | 41.76 | 41.61 | 41.55 | – | 42.61 | 42.79 | 5 | 43.81 | – | – | 43.99 | 42.66 | – | 3 |
| 20201216 | 19 | – | – | – | – | – | – | 0 | – | – | – | 42.73 | – | – | 1 |
| 20201217 | 20 | 41.49 | 41.63 | – | 42.80 | 42.05 | – | 4 | – | 43.96 | 42.62 | 43.82 | 41.92 | – | 4 |

"–": below limit of detection (Ct > 50).

**Table S5 (Continued).** Details of detection of SARS-CoV-2 RNA in wastewater of WWTP A.

| Date | Sample ID | Ct value for N1 (*n* = 6) | | | | | | Positive number  for N1 | Ct value for N2 (*n* = 6) | | | | | | Positive number  for N2 |
| --- | --- | --- | --- | --- | --- | --- | --- | --- | --- | --- | --- | --- | --- | --- | --- |
|  |  | 1 | 2 | 3 | 4 | 5 | 6 |  | 1 | 2 | 3 | 4 | 5 | 6 |  |
| 20201218 | 21 | 41.88 | 42.01 | 40.57 | 41.69 | 41.4 | 41.17 | 6 | – | 43.14 | 42.36 | 42.47 | 41.75 | 42.02 | 5 |
| 20201221 | 22 | 42.14 | 42.27 | 40.75 | 44.06 | 41.11 | 42.16 | 6 | 42.98 | 43.11 | 42.15 | 41.39 | 42.62 | 42.82 | 6 |
| 20201222 | 23 | 41.78 | 41.91 | 42.01 | 40.00 | 42.19 | 39.49 | 6 | 41.73 | 41.54 | 42.83 | 40.67 | – | 41.20 | 5 |
| 20201223 | 24 | 41.66 | 39.96 | 40.25 | 41.09 | 41.87 | 41.10 | 6 | 41.69 | – | – | 41.49 | 42.43 | – | 3 |
| 20201224 | 25 | 40.61 | 38.66 | 40.06 | 40.35 | 41.12 | 39.33 | 6 | 42.91 | 41.44 | 41.62 | 41.01 | 40.08 | 40.17 | 6 |
| 20201225 | 26 | 41.69 | 40.16 | 40.92 | 39.97 | 40.79 | 40.60 | 6 | 42.26 | 43.01 | 43.21 | 41.94 | 43.06 | 46.03 | 6 |
| 20201228 | 27 | 40.65 | – | 41.76 | – | – | – | 2 | 41.60 | 41.65 | 42.96 | – | 41.13 | 44.32 | 5 |
| 20201229 | 28 | 41.01 | 42.08 | 40.36 | – | – | – | 3 | 42.32 | 42.81 | – | – | – | 43.73 | 3 |
| 20201230 | 29 | 40.33 | 41.16 | – | 43.03 | 40.79 | – | 4 | – | 42.02 | 41.08 | – | 43.49 | 43.20 | 4 |
| 20201231 | 30 | 41.92 | 43.02 | 40.88 | 41.18 | 42.45 | – | 5 | 43.00 | 43.79 | – | 45.09 | – | 42.18 | 4 |
| 20210101 | 31 | – | 40.78 | 40.28 | 42.52 | – | 40.23 | 4 | 43.15 | 42.38 | 41.34 | 41.79 | 42.21 | 42.89 | 6 |
| 20210104 | 32 | 39.57 | 39.22 | 40.04 | 41.52 | 39.92 | 39.53 | 6 | 42.02 | 41.18 | 41.09 | 41.28 | 41.00 | 42.62 | 6 |
| 20210105 | 33 | 39.30 | 40.62 | 41.39 | 41.20 | 39.82 | 41.21 | 6 | 41.57 | 40.71 | 42.30 | 41.56 | 41.62 | 40.32 | 6 |
| 20210106 | 34 | 40.42 | 39.83 | 40.61 | 40.39 | 42.17 | 40.95 | 6 | 42.53 | 44.05 | 42.72 | – | 42.42 | 43.81 | 5 |
| 20210107 | 35 | 41.04 | 39.48 | 39.20 | 40.51 | 40.42 | 39.79 | 6 | 43.36 | 41.37 | 43.17 | 42.19 | 48.78 | 40.22 | 6 |
| 20210108 | 36 | 41.35 | 40.73 | 40.39 | 41.50 | 40.55 | 41.50 | 6 | 41.87 | 42.74 | 43.77 | 42.65 | 43.96 | 41.78 | 6 |
| 20210112 | 37 | 39.37 | 40.22 | 39.34 | 40.06 | 41.1 | – | 5 | 44.12 | 42.68 | 41.41 | 40.65 | 43.08 | 42.59 | 5 |
| 20210113 | 38 | 39.75 | 39.88 | 41.41 | – | 40.53 | 41.17 | 5 | 41.05 | 41.45 | 44.26 | – | 41.98 | 42.65 | 5 |
| 20210114 | 39 | 39.68 | 38.42 | 38.88 | 38.43 | 38.28 | 40.18 | 6 | 39.45 | 41.25 | 41.64 | 42.74 | 41.10 | 40.76 | 6 |
| 20210115 | 40 | 40.35 | – | 40.92 | – | 41.23 | – | 3 | 43.42 | – | 42.24 | – | 42.14 | 42.65 | 4 |

"–": below limit of detection (Ct > 50).

**Table S5 (Continued).** Details of detection of SARS-CoV-2 RNA in wastewater of WWTP A.

| Date | Sample ID | Ct value for N1 (*n* = 6) | | | | | | Positive number  for N1 | Ct value for N2 (*n* = 6) | | | | | | Positive number  for N2 |
| --- | --- | --- | --- | --- | --- | --- | --- | --- | --- | --- | --- | --- | --- | --- | --- |
|  |  | 1 | 2 | 3 | 4 | 5 | 6 |  | 1 | 2 | 3 | 4 | 5 | 6 |  |
| 20210118 | 41 | 41.24 | – | – | – | – | 41.91 | 2 | – | – | – | 42.44 | – | – | 1 |
| 20210119 | 42 | – | 41.45 | – | 41.68 | – | 41.54 | 3 | – | – | 44.91 | – | 43.29 | – | 2 |
| 20210120 | 43 | – | – | – | – | – | – | 0 | – | – | – | – | 42.29 | – | 1 |
| 20210121 | 44 | 39.81 | 39.72 | 41.23 | 40.87 | 40.93 | 41.68 | 6 | – | 44.32 | – | 41.03 | 43.43 | 46.03 | 4 |
| 20210122 | 45 | – | – | 41.52 | 41.92 | – | 41.52 | 3 | – | – | 43.86 | 45.08 | – | 44.34 | 3 |
| 20210125 | 46 | 39.27 | 39.40 | 38.88 | 38.78 | 40.57 | 39.76 | 6 | 43.09 | 49.13 | 42.98 | 40.94 | 41.92 | – | 5 |
| 20210126 | 47 | – | – | 42.10 | – | 42.32 | – | 2 | – | – | – | 42.31 | – | – | 1 |
| 20210127 | 48 | 41.08 | 41.07 | 40.22 | 42.1 | 41.30 | 39.84 | 6 | – | – | – | 42.59 | 45.6 | 43.70 | 3 |
| 20210128 | 49 | – | – | 40.34 | 39.87 | 40.17 | 41.29 | 4 | – | – | – | – | – | – | 0 |
| 20210129 | 50 | – | 44.08 | – | 42.37 | 41.52 | – | 3 | – | – | – | – | – | – | 0 |
| 20210201 | 51 | – | 41.26 | 41.23 | 40.48 | – | – | 3 | – | – | – | – | – | – | 0 |
| 20210202 | 52 | – | – | 41.79 | – | 40.96 | – | 2 | – | 43.14 | – | 44.49 | – | – | 2 |
| 20210203 | 53 | – | – | – | – | – | – | 0 | – | – | – | – | – | – | 0 |
| 20210204 | 54 | – | – | – | – | – | – | 0 | – | – | – | 42.62 | – | – | 1 |
| 20210205 | 55 | – | – | – | – | – | – | 0 | – | – | – | – | – | – | 0 |
| 20210208 | 56 | – | – | – | – | – | – | 0 | – | – | – | – | – | – | 0 |
| 20210209 | 57 | – | – | – | – | – | 43.99 | 1 | – | – | – | – | – | – | 0 |
| 20210210 | 58 | – | – | – | – | – | – | 0 | – | – | – | – | – | 42.71 | 1 |
| 20210212 | 59 | 48.72 | – | 44.32 | – | – | 41.75 | 3 | 43.07 | – | – | – | 43.16 | – | 2 |
| 20210215 | 60 | – | – | – | – | – | – | 0 | 43.80 | – | – | – | – | – | 1 |

"–": below limit of detection (Ct > 50).

**Table S6.** Comparison between epidemiological data of COVID-19 cases and wastewater-based epidemiology data.

| Date |  | Number of new cases in clinical test in the city B | | SARS-CoV-2 RNA in Wastewater | |
| --- | --- | --- | --- | --- | --- |
|  |  | by the date reported | by the date of symptom onset*^a^* | Wastewater sampling  (Sample ID)*^b^* | Total positive number for N1+N2*^c^* |
| 20201010 |  | 1 | 0 |  |  |
| 20201011 |  | 0 | 0 |  |  |
| 20201012 |  | 0 | 1 |  |  |
| 20201013 |  | 1 | 1 |  |  |
| 20201014 |  | 1 | 1 |  |  |
| 20201015 |  | 1 | 4 |  |  |
| 20201016 |  | 0 | 0 |  |  |
| 20201017 |  | 0 | 2 |  |  |
| 20201018 |  | 0 | 0 |  |  |
| 20201019 |  | 2 | 0 |  |  |
| 20201020 |  | 1 | 1 | 〇(1) | 0 |
| 20201021 |  | 1 | 0 |  |  |
| 20201022 |  | 1 | 0 |  |  |
| 20201023 |  | 1 | 1 | 〇(2) | 0 |
| 20201024 |  | 1 | 0 |  |  |
| 20201025 |  | 0 | 1 |  |  |
| 20201026 |  | 0 | 2 |  |  |
| 20201027 |  | 1 | 2 | 〇(3) | 0 |
| 20201028 |  | 2 | 3 |  |  |
| 20201029 |  | 0 | 1 |  |  |
| 20201030 |  | 0 | 0 | 〇(4) | 1 |
| 20201031 |  | 3 | 2 |  |  |

*a*: Some cases were confirmed positive without symptoms (asymptomatic cases). For these cases, the date of clinical test result reporting was counted as the estimated date of symptom onset. *b*: Circles indicate when wastewater samples were collected. IDs are from Table S5. *c*: Positive numbers in wastewater for each N1 and N2 assay are shown in Table S5.

**Table S6 (Continued).** Comparison between epidemiological data of COVID-19 cases and wastewater-based epidemiology data

| Date |  | Number of new cases in clinical test in the city B | | SARS-CoV-2 RNA in Wastewater | |
| --- | --- | --- | --- | --- | --- |
|  |  | by the date reported | by the date of symptom onset*^a^* | Wastewater sampling  (Sample ID)*^b^* | Total positive number for N1+N2*^c^* |
| 20201101 |  | 1 | 1 |  |  |
| 20201102 |  | 3 | 6 | 〇(5) | 3 |
| 20201103 |  | 1 | 0 |  |  |
| 20201104 |  | 0 | 3 |  |  |
| 20201105 |  | 8 | 2 |  |  |
| 20201106 |  | 1 | 2 | 〇(6) | 8 |
| 20201107 |  | 1 | 3 |  |  |
| 20201108 |  | 8 | 7 |  |  |
| 20201109 |  | 10 | 9 |  |  |
| 20201110 |  | 1 | 4 | 〇(7) | 6 |
| 20201111 |  | 4 | 5 |  |  |
| 20201112 |  | 4 | 2 |  |  |
| 20201113 |  | 0 | 6 | 〇(8) | 9 |
| 20201114 |  | 2 | 1 |  |  |
| 20201115 |  | 4 | 3 |  |  |
| 20201116 |  | 3 | 5 |  |  |
| 20201117 |  | 5 | 3 | 〇(9) | 4 |
| 20201118 |  | 1 | 3 |  |  |
| 20201119 |  | 3 | 3 |  |  |
| 20201120 |  | 5 | 3 | 〇(10) | 8 |
| 20201121 |  | 2 | 2 |  |  |
| 20201122 |  | 3 | 1 |  |  |

*a*: Some cases were confirmed positive without any symptom, i.e., asymptomatic cases. For these cases, the date of clinical test result reporting was counted as the estimated date of symptom onset. *b*: Circles indicate wastewater were collected at these dates. IDs are from Table S5. *c*: Positive numbers in wastewater for each N1 and N2 assay are shown in Table S5.

**Table S6 (Continued).** Comparison between epidemiological data of COVID-19 cases and wastewater-based epidemiology data

| Date |  | Number of new cases in clinical test in the city B | | SARS-CoV-2 RNA in Wastewater | |
| --- | --- | --- | --- | --- | --- |
|  |  | by the date reported | by the date of symptom onset*^a^* | Wastewater sampling  (Sample ID)*^b^* | Total positive number for N1+N2*^c^* |
| 20201123 |  | 1 | 0 |  |  |
| 20201124 |  | 2 | 2 | 〇 (11) | 1 |
| 20201125 |  | 3 | 1 |  |  |
| 20201126 |  | 1 | 2 |  |  |
| 20201127 |  | 1 | 1 | 〇 (12) | 2 |
| 20201128 |  | 1 | 1 |  |  |
| 20201129 |  | 0 | 1 |  |  |
| 20201130 |  | 1 | 1 |  |  |
| 20201201 |  | 3 | 1 | 〇 (13) | 0 |
| 20201202 |  | 2 | 2 |  |  |
| 20201203 |  | 1 | 1 |  |  |
| 20201204 |  | 1 | 1 | 〇 (14) | 5 |
| 20201205 |  | 0 | 2 |  |  |
| 20201206 |  | 0 | 1 |  |  |
| 20201207 |  | 3 | 1 |  |  |
| 20201208 |  | 2 | 1 | 〇 (15) | 2 |
| 20201209 |  | 0 | 2 |  |  |
| 20201210 |  | 2 | 2 |  |  |
| 20201211 |  | 4 | 1 | 〇 (16) | 4 |
| 20201212 |  | 0 | 3 |  |  |
| 20201213 |  | 4 | 9 |  |  |
| 20201214 |  | 3 | 6 | 〇 (17) | 11 |

*a*: Some cases were confirmed positive without any symptom, i.e., asymptomatic cases. For these cases, the date of clinical test result reporting was counted as the estimated date of symptom onset. *b*: Circles indicate wastewater were collected at these dates. IDs are from Table S5. *c*: Positive numbers in wastewater for each N1 and N2 assay are shown in Table S5.

**Table S6 (Continued).** Comparison between epidemiological data of COVID-19 cases and wastewater-based epidemiology data

| Date |  | Number of new cases in clinical test in the city B | | SARS-CoV-2 RNA in Wastewater | |
| --- | --- | --- | --- | --- | --- |
|  |  | by the date reported | by the date of symptom onset*^a^* | Wastewater sampling  (Sample ID)*^b^* | Total positive number for N1+N2*^c^* |
| 20201215 |  | 6 | 5 | 〇 (18) | 8 |
| 20201216 |  | 7 | 7 | 〇 (19) | 1 |
| 20201217 |  | 1 | 7 | 〇 (20) | 8 |
| 20201218 |  | 6 | 6 | 〇 (21) | 11 |
| 20201219 |  | 10 | 15 |  |  |
| 20201220 |  | 10 | 11 |  |  |
| 20201221 |  | 3 | 9 | 〇 (22) | 12 |
| 20201222 |  | 2 | 6 | 〇 (23) | 11 |
| 20201223 |  | 11 | 11 | 〇 (24) | 9 |
| 20201224 |  | 9 | 8 | 〇 (25) | 12 |
| 20201225 |  | 13 | 19 | 〇 (26) | 12 |
| 20201226 |  | 23 | 20 |  |  |
| 20201227 |  | 6 | 9 |  |  |
| 20201228 |  | 11 | 18 | 〇 (27) | 7 |
| 20201229 |  | 14 | 8 | 〇 (28) | 6 |
| 20201230 |  | 13 | 12 | 〇 (29) | 8 |
| 20201231 |  | 10 | 7 | 〇 (30) | 9 |
| 20210101 |  | 13 | 21 | 〇 (31) | 10 |
| 20210102 |  | 26 | 24 |  |  |
| 20210103 |  | 20 | 13 |  |  |
| 20210104 |  | 19 | 28 | 〇 (32) | 12 |
| 20210105 |  | 13 | 26 | 〇 (33) | 12 |

*a*: Some cases were confirmed positive without any symptom, i.e., asymptomatic cases. For these cases, the date of clinical test result reporting was counted as the estimated date of symptom onset. *b*: Circles indicate wastewater were collected at these dates. IDs are from Table S5. *c*: Positive numbers in wastewater for each N1 and N2 assay are shown in Table S5.

**Table S6 (Continued).** Comparison between epidemiological data of COVID-19 cases and wastewater-based epidemiology data

| Date |  | Number of new cases in clinical test in the city B | | SARS-CoV-2 RNA in Wastewater | |
| --- | --- | --- | --- | --- | --- |
|  |  | by the date reported | by the date of symptom onset*^a^* | Wastewater sampling  (Sample ID)*^b^* | Total positive number for N1+N2*^c^* |
| 20210106 |  | 21 | 14 | 〇 (34) | 11 |
| 20210107 |  | 22 | 13 | 〇 (35) | 12 |
| 20210108 |  | 14 | 18 | 〇 (36) | 12 |
| 20210109 |  | 15 | 12 |  |  |
| 20210110 |  | 11 | 13 |  |  |
| 20210111 |  | 13 | 8 |  |  |
| 20210112 |  | 4 | 12 | 〇 (37) | 10 |
| 20210113 |  | 13 | 6 | 〇 (38) | 10 |
| 20210114 |  | 19 | 14 | 〇 (39) | 12 |
| 20210115 |  | 9 | 7 | 〇 (40) | 7 |
| 20210116 |  | 13 | 12 |  |  |
| 20210117 |  | 8 | 7 |  |  |
| 20210118 |  | 3 | 10 | 〇 (41) | 3 |
| 20210119 |  | 9 | 10 | 〇 (42) | 5 |
| 20210120 |  | 11 | 6 | 〇 (43) | 1 |
| 20210121 |  | 9 | 9 | 〇 (44) | 10 |
| 20210122 |  | 11 | 8 | 〇 (45) | 6 |
| 20210123 |  | 12 | 9 |  |  |
| 20210124 |  | 7 | 12 |  |  |
| 20210125 |  | 5 | 8 | 〇 (46) | 11 |
| 20210126 |  | 16 | 8 | 〇 (47) | 3 |
| 20210127 |  | 4 | 5 | 〇 (48) | 9 |

*a*: Some cases were confirmed positive without any symptom, i.e., asymptomatic cases. For these cases, the date of clinical test result reporting was counted as the estimated date of symptom onset. *b*: Circles indicate wastewater were collected at these dates. IDs are from Table S5. *c*: Positive numbers in wastewater for each N1 and N2 assay are shown in Table S5.

**Table S6 (Continued).** Comparison between epidemiological data of COVID-19 cases and wastewater-based epidemiology data

| Date |  | Number of new cases in clinical test in the city B | | SARS-CoV-2 RNA in Wastewater | |
| --- | --- | --- | --- | --- | --- |
|  |  | by the date reported | by the date of symptom onset*^a^* | Wastewater sampling  (Sample ID)*^b^* | Total positive number for N1+N2*^c^* |
| 20210128 |  | 6 | 4 | 〇 (49) | 4 |
| 20210129 |  | 8 | 4 | 〇 (50) | 3 |
| 20210130 |  | 2 | 6 |  |  |
| 20210131 |  | 1 | 1 |  |  |
| 20210201 |  | 3 | 1 | 〇 (51) | 3 |
| 20210202 |  | 9 | 6 | 〇 (52) | 4 |
| 20210203 |  | 2 | 3 | 〇 (53) | 0 |
| 20210204 |  | 3 | 2 | 〇 (54) | 1 |
| 20210205 |  | 3 | 1 | 〇 (55) | 0 |
| 20210206 |  | 2 | 1 |  |  |
| 20210207 |  | 0 | 1 |  |  |
| 20210208 |  | 1 | 0 | 〇 (56) | 0 |
| 20210209 |  | 3 | 1 | 〇 (57) | 1 |
| 20210210 |  | 0 | 1 | 〇 (58) | 1 |
| 20210211 |  | 1 | 1 |  |  |
| 20210212 |  | 0 | 1 | 〇 (59) | 5 |
| 20210213 |  | 2 | 0 |  |  |
| 20210214 |  | 0 | 1 |  |  |
| 20210215 |  | 1 | 0 | 〇 (60) | 1 |
| 20210216 |  | 1 | 1 |  |  |
| 20210217 |  | 0 | 2 |  |  |
| 20210218 |  | 1 | 0 |  |  |

*a*: Some cases were confirmed positive without any symptom, i.e., asymptomatic cases. For these cases, the date of clinical test result reporting was counted as the estimated date of symptom onset. *b*: Circles indicate wastewater were collected at these dates. IDs are from Table S5. *c*: Positive numbers in wastewater for each N1 and N2 assay are shown in Table S5.

**Table S6 (Continued).** Comparison between epidemiological data of COVID-19 cases and wastewater-based epidemiology data

| Date |  | Number of new cases in clinical test in the city B | | SARS-CoV-2 RNA in Wastewater | |
| --- | --- | --- | --- | --- | --- |
|  |  | by the date reported | by the date of symptom onset*^a^* | Wastewater sampling  (Sample ID) | Total positive number for N1+N2*^c^* |
| 20210219 |  | 2 | 1 |  |  |
| 20210220 |  | 1 | 2 |  |  |
| 20210221 |  | 5 | 8 |  |  |
| 20210222 |  | 9 | 4 |  |  |
| 20210223 |  | 7 | 7 |  |  |
| 20210224 |  | 1 | 0 |  |  |
| 20210225 |  | 2 | 0 |  |  |

*a*: Some cases were confirmed positive without any symptom, i.e., asymptomatic cases. For these cases, the date of clinical test result reporting was counted as the estimated date of symptom onset.

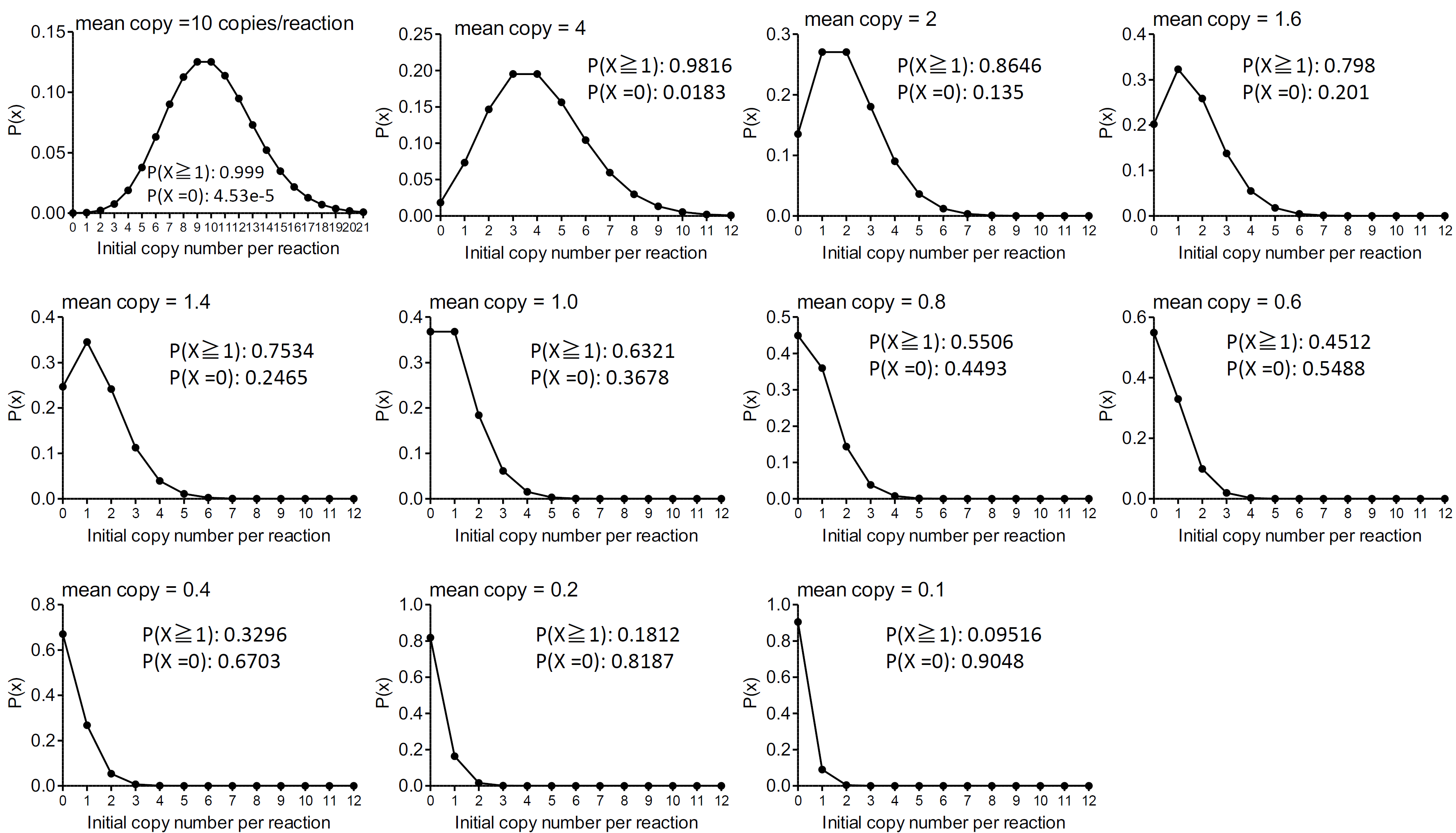

**Figure S1.** Poisson distribution of the probability of the number of molecules per reaction in qPCR. The figure shows the probability *P*(*x*) of obtaining a certain number of target molecules in a given volume when the initial copy number (*x*) is 10, 4, 2, 1.6, 1.4, 1.0, 0.8, 0.6, 0.4, 0.2, or 0.1 target molecules per reaction. The expected probabilities of a positive (*P*(*x* ≥ 1)) and negative reaction (*P*(*x* = 0)) by qPCR are shown for each case.

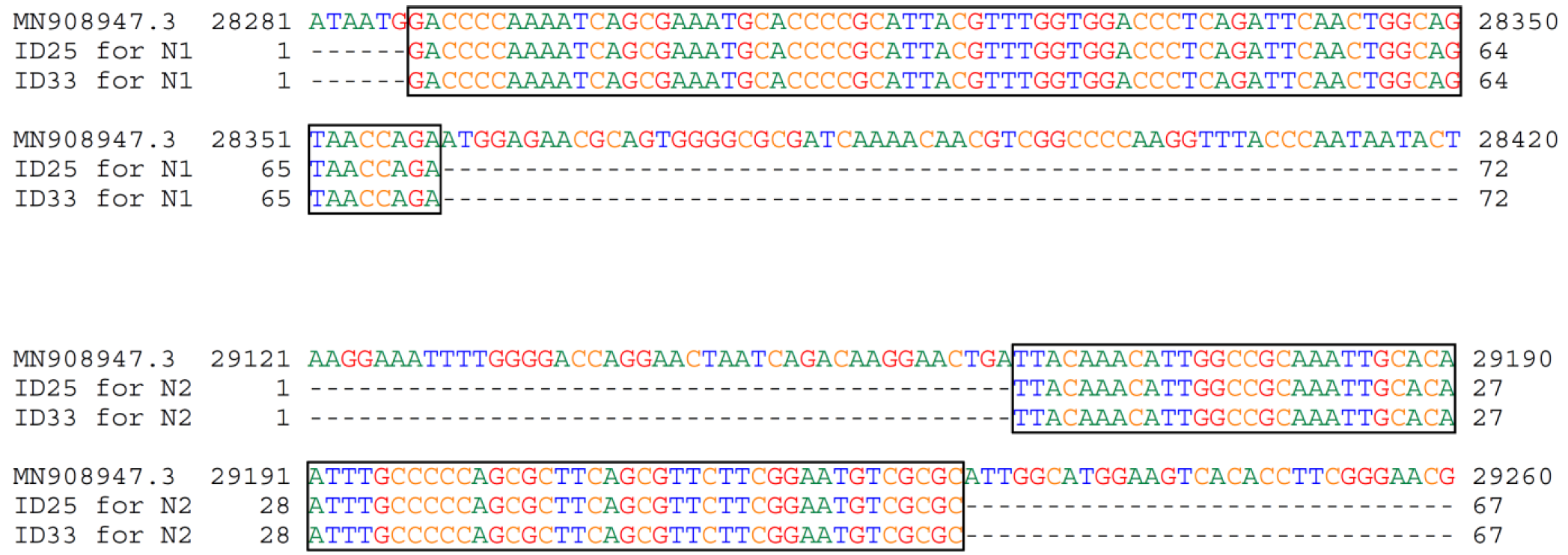

**Figure S2.** Sequences of qPCR products of wastewater samples ID25 and ID33 by N1 (top, 72 bp) and N2 assays (bottom, 67 bp) were aligned to the SARS-CoV-2 genome (GenBank Acc. No. MN908947.3).

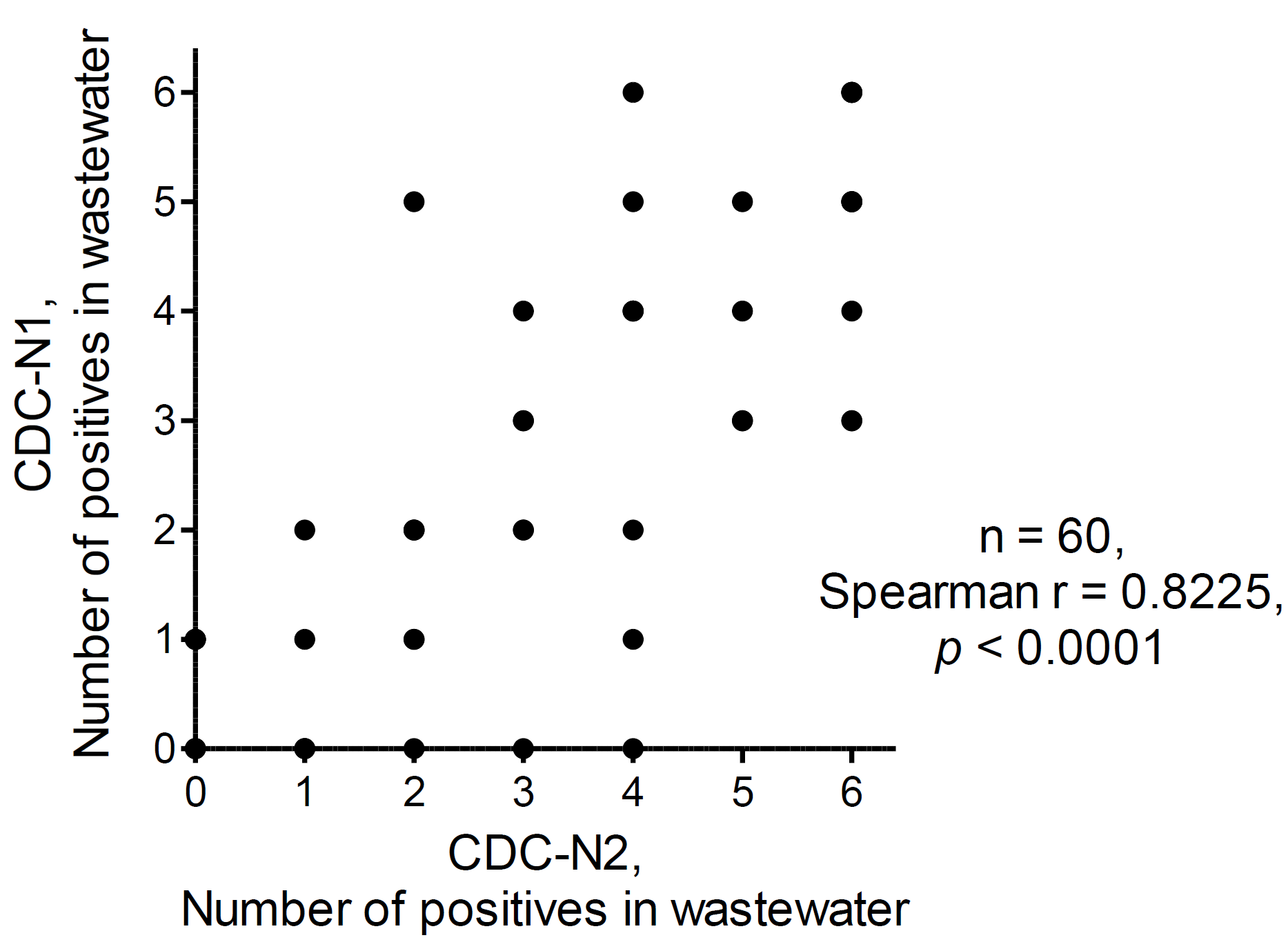

**Figure S3.** Correlations of numbers of positive reactions between CDC-N1 and CDC-N2 assays of wastewater from WWTP A by Spearman's rank correlation test (*n* = 60).

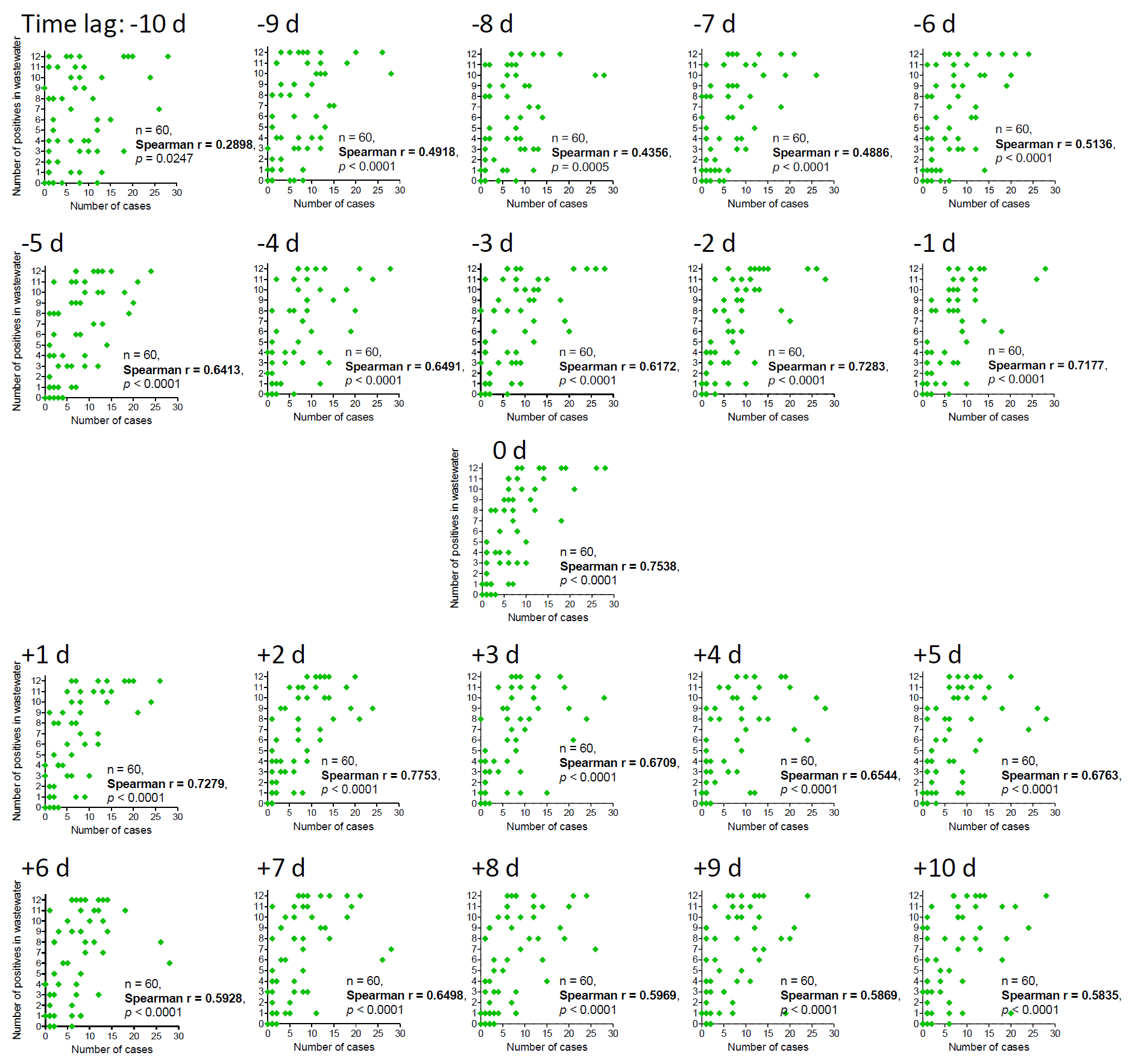

**Figure S4.** Time-step analysis of correlations between new cases by clinical testing and positive numbers by N1 + N2 assays in wastewater by Spearman's rank correlation test. SARS-CoV-2 RNA signals in wastewater were compared with the numbers of new cases at the wastewater sampling day (time lag = 0); from 10 days to 1 day before (time lag = −10 to −1); and from 1 to 10 days after the wastewater sampling day (time lag = +1 to +10).
